# Safety and effectiveness of dose-sparing strategies for seasonal influenza vaccine

**DOI:** 10.1101/2020.07.31.20163717

**Authors:** Jesmin Antony, Patricia Rios, Chantal Williams, Naveeta Ramkissoon, Sharon E. Straus, Andrea C. Tricco

## Abstract

**Background:** The objective of this rapid scoping review was to identify potentially safe and effective dose-sparing strategies for intramuscular administration of seasonal influenza vaccines in healthy individuals of all ages.

**Methods:** Comprehensive literature searches were developed and executed in MEDLINE, EMBASE, and the Cochrane library, and grey literature was searched via international clinical trial registries for relevant studies published in English in the last 20 years. References of relevant systematic reviews and included studies were also scanned. Title/abstract and full-text screening were carried out by pairs of reviewers independently and data charting conducted by a single reviewer and verified by a second reviewer. Results were presented narratively.

**Results:** A total of 13 studies with 10,351 participants were included in the review and all studies were randomized control trials conducted between 2006 and 2019. The most common interventions were the trivalent influenza vaccine (n=10), followed by quadrivalent influenza vaccine (n=4). Nine studies included infants/toddlers 6-36 months old and one of these studies also included children and adolescents. In these nine studies, no clinical effectiveness outcomes were reported and no difference was found in local and systemic reactogenicity between dosing strategies. Of the four adult studies (≥ 18 years), the two studies that reported on effectiveness outcomes found similar results between the half-dose and full-dose vaccination groups and all four studies reported no differences in safety outcomes between groups.

**Conclusion:** The current evidence for the administration of intramuscular influenza vaccines suggests there is no significant difference in safety and clinical effectiveness with the use of low-dose compared to full-dose vaccines, which is promising given the predicted resource constraints in the upcoming influenza season due to the 2019 novel coronavirus. Due to the low number of studies in adults and the lack of studies assessing confirmed influenza and influenza-like illness, there remains a need for further evaluation.

**EXECUTIVE SUMMARY:** *PURPOSE:* The Centre for Immunization and Respiratory Infectious Diseases of the Public Health Agency of Canada (PHAC) submitted a query regarding the safety and effectiveness of fractional dosing of seasonal influenza vaccines through the Canadian Institutes of Health Research (CIHR) Drug Safety and Effectiveness Network (DSEN). They requested that the DSEN Methods and Application Group in Indirect Comparisons (MAGIC) conduct a rapid review on this topic with an approximate 6-week timeline. The overall objective of this rapid review was to identify potentially safe and effective dose-sparing strategies for administration of seasonal influenza vaccines in healthy individuals of all ages that have been evaluated in human trials. In order to limit the scope of the work and ensure the rapid timeline could be met, this review focused only on intramuscular vaccine formulations, thus the research question was as follows: 1. What is the safety and effectiveness of using fractional dosing strategies to deliver intramuscular seasonal influenza vaccines?

## METHODS

### Protocol

The methods for this review were guided by the updated reviewer manual published by the Joanna Briggs Institute and the World Health Organization’s guide to rapid evidence synthesis.^1, 2^ Results are reported according to the Preferred Reporting Items for Systematic Reviews and Meta-analysis extension to scoping reviews (PRISMA-ScR).^3^ A protocol for this rapid review was published on the Open Science Framework registry (https://osf.io/8mwz2/).

### Literature search

Comprehensive literature searches were developed and executed in MEDLINE (available in Appendix A), EMBASE, and the Cochrane library, and grey (i.e., difficult to locate or unpublished) literature was searched via international clinical trial registries. References of relevant systematic reviews and included studies were also scanned.

### Eligibility criteria

The eligibility criteria followed the PICOST framework:

- <u>Population:</u> Healthy humans of any age. Immunocompromised populations and animal studies were excluded.
- <u>Intervention:</u> Any dose-sparing strategy used to administer intramuscular seasonal influenza vaccines (vaccines of interest listed in Appendix B). Eligible strategies include, but were not limited to, administrating less than the standard 15 ug HA antigen using multi-dose vials, half dosing, or pre-formulated products with reduced antigen quantity, or using revised vaccine schedules to distribute doses. Any studies examining monovalent pandemic vaccines, specialty/experimental vaccines (e.g., high dose), whole virus vaccines, or other routes of administration (e.g. intranasal, intradermal) were not eligible. Only vaccine products approved for use in Canada or equivalent formulations approved for use in other countries were eligible for inclusion. Concomitant administration with other vaccine products were included only if administered to both the intervention and the comparator groups.
- <u>Comparator:</u> Any of the interventions listed above, no intervention, or placebo.
- <u>Outcomes:</u> Lab-confirmed influenza infection (primary outcome), influenza-like illness or clinical/symptomatic diagnosis of influenza, hospitalization, ICU admission, pneumonia, mortality, and adverse events (local/systemic reactogenicity, vascular-related, serious).
- <u>Study designs:</u> Randomized controlled trials (RCTs), NRCTs (e.g., such as quasi-RCTs, non-randomized trials, interrupted time series, controlled before after), and observational studies (e.g., cohort, case control) were included. Studies must have a control or comparator in order to be eligible for inclusion and as such, cross-sectional, case series, case reports, and qualitative studies were excluded.
- <u>Time periods:</u> Only studies published in the past 20 years (2000-2020) were included.

Inclusion was also limited to studies written in the English language only due to the short timelines for this review.

### Study selection

A screening form based on the eligibility criteria was prepared and pilot-tested with all members of the review team until sufficient agreement (≥75%) was reached prior to both title/abstract (level 1) and full-text (level 2) screening. Subsequent screening at level 1 and level 2 were completed by pairs of reviewers working independently using the Knowledge Translation Program’s proprietary screening software (synthesi.SR).^4^ Any discrepancies between reviewers were resolved by a third independent reviewer.

### Data items and charting process

Items for data collection included study characteristics (e.g., study design, year of publication, country of conduct, multi-center vs. single site), patient characteristics (e.g., mean age, age range, sex, vaccination history), intervention details (e.g., type of vaccine, vaccine manufacturer, dose, timing an administration of treatment), comparator details (e.g., comparator intervention, dose), and outcome results (e.g., influenza infections, hospitalizations, adverse events, mortality) at the longest duration of follow-up. Immunogenicity outcomes were not abstracted, but these studies were flagged for PHAC.

A standardized form for data charting was developed and pilot tested by the entire review team using 2 full-text articles to ensure congruence among reviewers. All included studies were charted by one reviewer and then verified by a second reviewer working independently.

## RESULTS

### Literature search

We screened 2378 titles and abstracts from our database search and an additional 13 citations located through searching the grey literature and scanning references. Of these, 144 potentially relevant full-text articles were screened for eligibility and data from 13 relevant studies were abstracted. Five trial protocols related to these 13 full-text articles were also captured in our search and have been denoted as companion reports (Figure 1). Twelve studies that assessed dose-sparing strategies were excluded during full-text screening because the vaccine under study was not of interest or unclear. We contacted authors of the unclear studies and received 1 response confirming the vaccine was not of interest. These 12 studies are listed in Appendix C.

**Figure 1:**
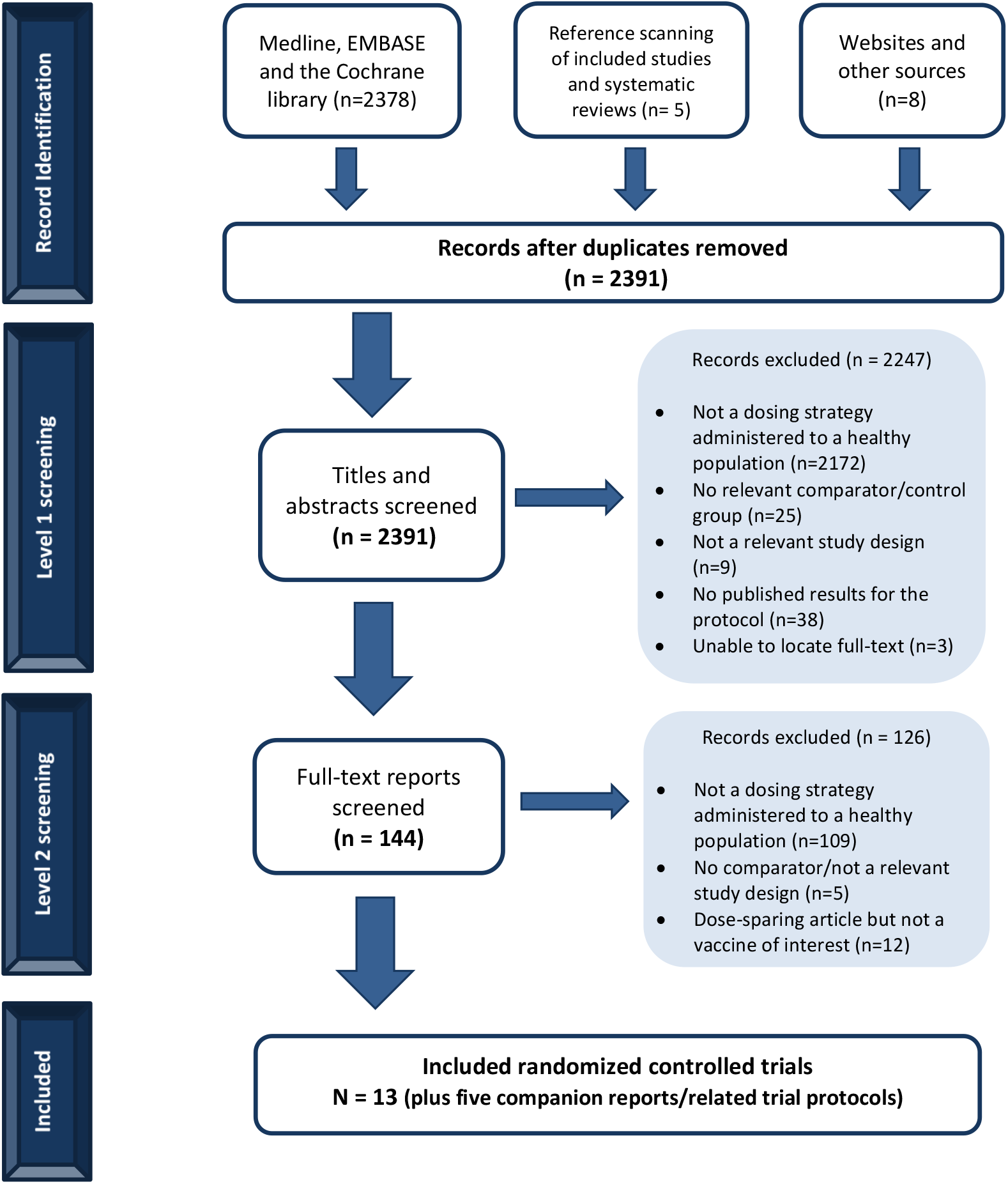
Study Flow Diagram

### Study characteristics

Table 1 summarizes the characteristics of the 13 included studies. All studies were randomized controlled trials conducted between 2006 and 2019; mainly in the US, followed by Mexico, Canada and Finland. The majority of the studies evaluated trivalent vaccines (77%) and most were conducted in the 6-36 month-old pediatric population (69%). Almost all studies reported on reactogenicity and/or adverse events, but only two studies reported on effectiveness outcomes of interest (i.e., confirmed influenza and influenza-like illness).

**Table 1:** Summary of included studies.

|  |  | N (%) |
| --- | --- | --- |
| **Total # of included studies** |  | **13 (100)** |
| **Date of publication** | 2006-2010 | 4 (30.8) |
|  | 2011-2015 | 5 (38.4) |
|  | 2016-2020 | 4 (30.8) |
| **Country(ies) of conduct<sup>a</sup>** | USA | 8 (61.5) |
|  | Mexico | 3 (23.1) |
|  | Canada | 2 (15.4) |
|  | Finland | 2 (15.4) |
|  | Belgium | 1 (7.69) |
|  | Hong Kong | 1 (7.69) |
|  | Taiwan | 1 (7.69) |
|  | Thailand | 1 (7.69) |
| **Study design** | Randomized controlled trial (RCT) | 13 (100%) |
| **Populations<sup>a,b</sup>** | Infants/Toddlers (6-36 months) | 9 (69.2) |
|  | Children (37 months – 17 years) | 1 (7.69) |
|  | Adults (18-64 years) | 3 (23.1) |
|  | Older adults (≥65) | 2 (15.4) |
| **Treatments<sup>a,c</sup>** | Trivalent influenza vaccine (TIV) | 10 (76.9) |
|  | Quadrivalent influenza vaccine (QIV) | 4 (30.8) |
| **Outcomes<sup>a</sup>** | Effectiveness | 2 (15.4) |
|  | Local and Systemic Reactogenicity | 12 (92.3) |
|  | Adverse events | 10 (76.9) |
|  | Immunogenicity | 12 (92.3) |

<sup>a</sup>Each study can fit into more than one category so the total percentage will not add up to 100%
<sup>b</sup>One study includes both infants/toddlers and children, and another includes both adults and seniors
<sup>c</sup>One study includes both TIV and QIV arms

Full study and patient characteristic details for each study are reported in Appendix C and treatment and outcome details in Appendix D.

### Studies including children (<18 years old)

Nine studies included infants/toddlers 6-36 months old and one study also included children and adolescents (Table 2). None of these studies reported results on the effectiveness outcomes established a priori, however all of them reported on safety outcomes. Immunogenicity outcomes were also reported in all these studies and flagged for PHAC in Table 2.

**Table 2:** Nine RCTs conducted in children (6 months – 17 years)

| Author, Year | Study period and countr(ies) | Treatment arms<br>Brand name (manufacturer)<br>HA/strain [dosing] | Mean age (SD) | ITT sample size | Relevant outcomes | Conclusions<br>[other outcomes reported but not abstracted] |
| --- | --- | --- | --- | --- | --- | --- |
| TRIVALENT AND QUADRIVALENT INFLUENZA VACCINES (TIV/QIV) |  |  |  |  |  |  |
| Cioppa, 2011<sup>5</sup> | October 2008 – March 2009<br>Belgium | NR - TIV, **7.5-µg/strain** [2 x 0.25mL dose] | 20.0 months (7.0) | 25 | Local and Systemic reactogenicity<br>Adverse events | Reactogenicity of the 7.5-µg TIV/QIV formulations was slightly lower than for the corresponding 15-µg formulations. |
|  |  | Agrippal - TIV, **15-µg/strain** [2 x 0.5mL dose] | 15.0 months (8.8) | 22 |  | The majority of unsolicited AEs were mild or moderate in severity and none of the SAEs was considered to be related to the study vaccine. |
|  |  | NR - QIV, **7.5-µg/strain** [2 x 0.25mL dose] | 18.0 months (8.9) | 25 |  |  |
|  |  | NR - QIV, **15-µg/strain** [2 x 0.5mL dose] | 15.2 months (7.8) | 28 |  |  |
|  |  | Vaxigrip (Sanofi Pasteur), **7.5-µg/strain** [2 x 0.25mL dose] | 16.1 months (8.5) | 26 |  | [Immunogenicity, Seroconversion, Sero-protection, GMT] |
| TRIVALENT INFLUENZA VACCINES (TIV) |  |  |  |  |  |  |
| Skowronski, 2011<sup>6</sup> | September 2008 – December 2008<br>Canada | Vaxigrip (Sanofi-Pasteur), 15-µg/strain [**2 x 0.5mL dose**] | 13.2 months (5.1) | 124 | Local and Systemic reactogenicity<br>Adverse events | Local reactions generally were less common in infants than toddlers and more common with full doses versus half doses, but none of these differences were significant. |
|  |  | Vaxigrip (Sanofi-Pasteur), 15-µg/strain [**2 x 0.25mL dose**] | 12.8 months (5.0) | 128 |  | One serious adverse event was reported: a toddler in the half dose group was hospitalized with pneumonia 28 days after the first vaccination. The event was deemed unlikely related to the vaccine. |
|  |  |  |  |  |  | Compared with 0.25-mL half-dosing, administration of 2 full 0.5-mL doses of trivalent inactivated influenza vaccine can increase antibody response without increasing reactogenicity in previously unimmunized infants aged 6 to 11 months. |
|  |  |  |  |  |  | [Immunogenicity, Sero-protection, Seroconversion] |
| Langley, 2012<sup>7</sup> | November 2008 – | Fluviral F1 (Sanofi-Pasteur), **7.5-µg/strain** [1 x 0.25 mL dose] | 18.2 months (9.06) | 164 | Local and Systemic reactogenicity | Fluviral F1 group had 1 case of pneumonia resolved. Fluviral F2 group had 1 case of bronchial hyper-reactivity in resolving stage. |
|  | August 2009<br>Canada | Fluviral F2 (Sanofi-Pasteur),<br>**15-µg/strain** [1 x 0.5mL dose] | 17.5 months (8.27) | 167 | Adverse events | The 0.5-mL dose of the study vaccine, when administered to children aged 6–35 months, resulted in a modest but not statistically significant improvement in immunogenicity with clinically similar safety and reactogenicity compared with the 0.25-mL dose.<br>[Immunogenicity, Seroconversion rate; Seroprotection rate] |
|  |  | Vaxigrip (Sanofi-Pasteur),<br>**7.5-µg/strain** [1 x 0.25 mL dose] | 17.0 months (8.33) | 43 |  |  |
| Pavia-Ruz, 2013<sup>8</sup> | October 2008-March 2009 | Fluarix (GSK),<br>**15-µg/strain** [1 x 0.5mL dose] | 21.2 months (8.37) | 1018 | Local and Systemic reactogenicity<br>Adverse events | The reactogenicity and safety profile of the study vaccine did not appear to be affected by doubling the dose.<br>One participant in the Flu-15µg group had two SAEs, (apnea and cyanosis) which were considered by the investigator to be possibly related to vaccination. The subject was hospitalized and the events resolved on the same day as they occurred.<br>[Immunogenicity, Seroconversion rate, Seroprotection rate, GMT, GMFR] |
|  | Hong Kong, Mexico, Taiwan, Thailand, and the USA | Fluarix (GSK),<br>**7.5-µg/strain** [1 x 0.25 mL dose] | 21.2 months (8.03) | 1018 |  |  |
|  |  | Fluzone (Sanofi-Pasteur),<br>**7.5-µg/strain** [1 x 0.25 mL dose] | 21.1 months (8.20) | 1031 |  |  |
| Halasa, 2015<sup>9</sup> | 2010-2012<br>USA | Fluzone (Sanofi Pasteur),<br>**7.5-µg/strain** [1 x 0.25 mL dose] | 13.5 | 80 | Local and Systemic reactogenicity | No significant differences between the full-dose or half-dose groups for either the fully primed or naive cohorts for systemic reactions or local reactions when both seasons were combined.<br>The only significant difference in the 2011–2012 season was that 8 of 48 (16.7%) participants in the half-dose group compared with 32 of 96 (33.3%) in the full-dose group had increased redness at the injection site (P < .05).<br>No significant differences between the groups in AE, SAE, or onset of chronic medical conditions between the dose groups in either the naive or fully primed cohorts, and none of the SAEs were deemed related to the vaccine.<br>[Immunogenicity, Seroprotection rate, HAI titers] |
|  |  | Fluzone (Sanofi Pasteur),<br>**15-µg/strain** [1 x 0.5 mL dose] | 14.5 | 163 |  |  |
| Phung, 2016<sup>10</sup> | September 2010-January 2011 | FLUAD (NR),<br>NR [1 x 0.5mL dose] | 68.7 months (18) | 60 | Local and Systemic reactogenicity | *Trial protocol with no author conclusions.* |
|  | Finland | FLUAD (NR),<br>NR [1 x 0.25 mL dose] | 60.4 months (23.2) | 75 | Adverse events |  |
|  |  | Agrippal S1 (NR),<br>NR [1 x 0.5mL dose] | 68 months (17.1) | 51 |  |  |
|  |  | Agrippal S1 (NR),<br>NR [1 x 0.25mL dose] | 32.4 months (1.9) | 11 |  | [Immunogenicity, GMR, GMT, Seroconversion rate, Seroprotection rate] |
| **QUADRIVALENT INFLUENZA VACCINES (QIV)** |  |  |  |  |  |  |
| **Jain, 2017<sup>11</sup>** | 2014-2015 Influenza Season | Flulaval (GSK),<br>**15-µg/strain** [1 x 0.5mL dose] | 19.7 months (8.7) | 1013 | Local and Systemic reactogenicity | None of the febrile seizures or the SAEs were considered by the investigator to be related to vaccination. |
|  | USA and New Mexico | Fluzone (Sanofi Pasteur),<br>**7.5-µg/strain** [1 x 0.25 mL dose] | 19.9 months (8.9) | 1028 | Adverse events | Double-dose vaccines may improve protection against influenza B in some young children and simplifies annual influenza vaccination by allowing the same vaccine dose to be used for all eligible children and adults.<br>[Immunogenicity, Seroconversion rates, Seroprotection rates, GMT] |
| **Ojeda, 2019<sup>12</sup>** | December 2017- January 2018 | Vaxigrip Tetra (Sanofi Pasteur)<br>**PFS 15-µg/strain** [1 x 0.5mL dose] | NR (6 months – 17 years) | 149 | Local and Systemic reactogenicity | Solicited systemic reactions were reported in more infants aged 6 – 35 months in the MDV group than in the PFS group however this was not clinically significant. |
|  | Mexico | Vaxigrip Tetra (Sanofi Pasteur)<br>**MDV 15-µg/strain** [1 x 0.5mL dose] | NR (6 months – 17 years) | 153 | Adverse events | AE not considered related to a study vaccine.<br>There were no differences in reactogenicity or safety between the two vaccine formats. These results showed that the MDV format of QIV was as safe and immunogenic as the PFS format in infants, children, and adolescents. These findings support the use of MDV QIV as a resource-saving alternative for seasonal influenza vaccination.<br>[Immunogenicity, Seroconversion rates, HAI titers, GMT ratios] |
| **Robertson, 2019<sup>13</sup>** | September 2016 – March 2017<br>USA | Fluzone (Sanofi Pasteur)<br>**15-µg/strain** [1x0.5mL dose] | 20.5 months (8.55) | 992 | Local and Systemic reactogenicity | No significant differences between full- and half-dose groups.<br>AE leading to study discontinuation/SAE not considered vaccine-related. |
|  |  | Fluzone (Sanofi Pasteur)<br>**7.5-µg/strain** [1x0.25 dose] | 20.4 months (8.75) | 949 | Adverse events | A full dose vaccine was immunogenic and had a safety profile comparable to that of a half dose, with no new safety concerns observed.<br>[Immunogenicity, seroconversion rate] |

**Abbreviations:** AE – adverse events; GMR - geometric mean ratio; GMFR – geometric mean fold rise; GMT - geometric mean antibody titer; HA - hemagglutinin; HAI - hemagglutination inhibition; ID – intradermal; IM – intramuscular; ITT – intent-to-treat; MDV – multi-dose vials, n – number of people with condition, N – sample size of treatment arm, NR – not reported, PFS – prefilled dose, SAE – serious adverse events

### Safety outcomes

#### Trivalent influenza vaccines

Six of the included studies assessed trivalent influenza vaccines (TIV) in young children (6-36 months) and reported on local and systemic reactogenicity outcomes and adverse events.^5-10^ Two studies compared the administration of full- (0.5mL) and half- (0.25mL) doses of the same standard 15μg/strain vaccine.^6, 10^ The first RCT compared 2 full versus 2 half doses of TIV in previously unimmunized infants (6-11 months) and toddlers (12-23 months) using Vaxigrip (15μg/strain).^6^ The study found that in the infants group, 2 full 0.5-mL doses of vaccine did not increase reactogenicity. Local reactions were less common in infants than toddlers and more common with full doses versus half doses, but none of these differences were statistically significant. All adverse events reported in the study were deemed unlikely related to the vaccine. The second study, published in a clinical trial registry, compared a single intramuscular injection of 0.5mL to 0.25mL of FLUAD or Agrippal and showed comparable proportions of children with reactogenicity outcomes and AEs across the groups, but no significance levels or conclusions were provided by the investigators.^10^

The objective of three of the included trials was to examine the impact of administering the full adult dose of 15μg/strain vaccines compared with the usual children’s dose of 7.5μg/strain in infants and toddlers.^7-9^ A multicenter randomized trial was conducted in Canada assessing the safety of full-dose Fluviral TIV (15μg/strain) compared with the half-dose (7.5μg/strain) and an active comparator Vaxigrip (7.5μg/strain).^7^ Compared with the half-dose, the full-dose of the study vaccine resulted in clinically similar reactogenicity and safety. A similar three-arm randomized study to assess the use of Fluarix at two different dose levels (7.5μg/strain and 15μg/strain) compared to an established control vaccine Fluzone (7.5μg/strain) also found the reactogenicity and safety profile of Fluarix did not appear to be affected by doubling the dose, but one participant in the 15μg group had two SAEs (apnea and cyanosis) that were considered by the investigator to be possibly related to vaccination.^8^ A third multicenter trial compared the 15 μg/strain formulation to the 7.5μg/strain formulation of Fluzone (Sanofi Pasteur) administered to young children across multiple influenza seasons.^9^ This study also found no statistically significant differences between the full-dose or half-dose groups for systemic reactions, local reactions or adverse events when both seasons were combined; however, in the 2011–2012 season, 8 of 48 (16.7%) participants in the half-dose group compared with 32 of 96 (33.3%) in the full-dose group had increased redness at the injection site (P < .05).

Cioppa et al. (2009) was the only trial that compared the safety and tolerability of both TIV and QIV vaccine formulations.^5^ The vaccine arms of interest were a QIV 15-μg/strain, TIV 15-μg/strain, QIV 7.5-μg/strain, TIV 7.5-μg/strain, and a control Vaxigrip TIV 7.5-μg/strain vaccine. Reactogenicity of the 7.5-μg TIV/QIV formulations was slightly lower than for the corresponding 15-μg formulations, but there was no difference in reactogenicity between TIV and QIV vaccines.

#### Quadrivalent influenza vaccines

Four of the included studies evaluated quadrivalent influenza vaccines (QIV) in children.^5, 11-13^All of the studies reported reactogenicity outcomes and adverse events. One study reported both TIV and QIV vaccines and the results are reported above.^5^ Two studies compared full-dose QIV to pediatric 7.5μg/strain Fluzone. In the first trial, full dose Fluzone had a similar safety profile to half-dose Fluzone with a single adverse event being attributed to the study vaccine.^13^ Similarly, the second study found that full-dose Flulaval may improve protection against influenza in some young children when compared to low-dose Fluzone, and in this trial none of the adverse events were considered to be study-related by the investigator.^11^ The final trial evaluated Vaxigrip Tetra (15μg/strain) administered to children and adolescents in two different formats.^12^ Vaxigrip administered as a single dose using a pre-filled syringe (PFS) was compared to a 10-dose multi-dose vial (MDV). Systemic reactions were reported in more infants aged 6 − 35 months in the MDV group than in the PFS group, however this difference was not clinically significant. The authors concluded that there was no difference in reactogenicity or safety between the two vaccine formats in infants, children, and adolescents.

### Studies including adults (≥18 years old)

One study included adults over 18 years, 2 studies included adults from18-45 and 18-65 years old, and 1 study included older adults (≥ 65 years) (Table 3). Two studies reported on effectiveness outcomes and three on reactogenicity and adverse events. Immunogenicity outcomes were also reported in 3 studies and flagged for PHAC in Table 3. All 4 trials evaluated Fluzone QIV.

**Table 3:** Four RCTs conducted in adults (≥18 years old)

| Author, Year | Study period and countr(ies) | Treatment arms<br>Brand name (manufacturer)<br>HA/strain [dosing] | Mean age (SD) | ITT sample size | Relevant Outcomes | Conclusions<br>[other outcomes reported but not abstracted] |
| --- | --- | --- | --- | --- | --- | --- |
| **QUADRIVALENT INFLUENZA VACCINES (QIV)** |  |  |  |  |  |  |
| **Kramer, 2006<sup>14</sup>** | October 2004 – November 2004<br>USA | Fluzone (Aventis Pasteur),<br>**15-µg/strain** [1 x 0.5mL dose] | NR (>18 years) | 222 | Lab-confirmed influenza | There was no significant difference between the full-dose and half-dose groups in the diagnosis of influenza or in the proportion of participants self-reporting four or more symptoms consistent with influenza-like illness.<br>No adverse events were noted by participants from either group or reported to the IRB during the course of the study<br>[None] |
|  |  | Fluzone (Aventis Pasteur),<br>**7.5-µg/strain** [1 x 0.25 mL dose] | NR (>18 years) | 222 | Influenza-like illness<br>Adverse events |  |
| **Engler, 2008<sup>15</sup>** | November 2004 – December 2004<br>USA | Fluzone (Aventis Pasteur),<br>**15-µg/strain** [1 x 0.5mL dose] | NR (18 – 65 years) | 554 | Influenza-like illness<br>Hospital/ER visits | The relative risk of medical visits and hospitalizations for influenza-like illnesses were similar in the half- and full-dose group regardless of age, and there was no evidence of ILI symptom differences by sex or dose during the 21 days after immunizations.<br>Although injection site pain was greater for full- vs half-dose (19.9% vs 14.4%; p=.01), when analyzed for clinically significant |
|  |  | Fluzone (Aventis Pasteur),<br>**7.5-µg/strain** [1 x 0.25 mL dose] | NR<br>(18 – 65 years) | 556 | Local and Systemic reactogenicity<br>Adverse events | pain levels significant dose-dependent pain differences were not identified.<br>Joint and/or muscle pain were significantly different (p=.02 and p=.03, respectively) by dose.<br>No other adverse event differed significantly by dose.<br>[Immunogenicity (antibody)] |
| **Belshe, 2007<sup>16</sup>** | NR<br>USA | Fluzone (Sanofi-Pasteur),<br>**15-µg/strain** [1 x 0.5mL dose] | 31.5 years (9.6) | 31 | Local and Systemic reactogenicity | Intradermal (ID) vaccine induced significantly more local inflammatory response than Intramuscular (IM) vaccine but this did not translate into an increased immune response for ID vaccines compared to IM (primary comparison of this study was ID vs IM doses)<br>[Immunogenicity, Seroconversion] |
|  |  | Fluzone (Sanofi-Pasteur),<br>**9-µg/strain** [1 x 0.3mL dose] | 31.2 years (9.4) | 32 |  |  |
|  |  | Fluzone (Sanofi-Pasteur),<br>**6-µg/strain** [1 x 0.2mL dose] | 30.1 years (10.3) | 31 |  |  |
|  |  | Fluzone (Sanofi-Pasteur),<br>**3-µg/strain** [1 x 0.1mL dose] | 31.9 years (10.3) | 31 |  |  |
| **Chi, 2010<sup>17</sup>** | August 2007-2008 | Fluzone (Sanofi Pasteur),<br>**15-µg/strain** [1 x 0.5mL dose] | 75.6 years (6.8) | 65 | Local and Systemic reactogenicity | The two SAEs were acute coronary syndrome and appendicitis and neither were judged to be related to influenza vaccination<br>[Immunogenicity, Seroprotection, GMT] |
|  | USA | Fluzone (Sanofi Pasteur),<br>**9-µg/strain** [1 x 0.3mL dose] | 75.2 years (7.7) | 64 | Adverse events |  |

**Abbreviations:** AE – adverse events, GMT - geometric mean antibody titer; HA - hemagglutinin; ID – intradermal; ILI – influenza-like illness; IM – intramuscular; MDV – multi-dose vials, n – number of people with condition, N – sample size of treatment arm, NR – not reported, PFS – prefilled syringe, SAE – serious adverse events

### Effectiveness outcomes

Two of the included studies that examined the same vaccine (Fluzone manufactured by Aventis Pasteur) in adult populations reported effectiveness outcomes including lab-confirmed influenza infections, influenza like illness, and/or hospitalizations or emergency room visits after vaccination.^14, 15^ The study by Kramer et al. (2006) found that 3.6% of participants receiving a 15-μg/strain dose of vaccine reported influenza like illness compared to 6.8% of participants that received a 7.5-μg/strain dose.^14^ However, only one participant in the study that received the 15-μg/strain dose was confirmed via laboratory analysis to have influenza. The authors concluded that half-dose and full-dose vaccinations appear to be similarly effective based on the low rate of influenza infections and similar symptom surveys between both groups but acknowledge that further studies examining immunogenicity are needed to confirm.

A similar study by Engler et al. (2008) that compared a 15-μg/strain dose of Fluzone vaccine to a 7.5-μg/strain dose found equal proportions of participants reporting influenza like illness (9.7% vs 9.9%) and hospitalizations or emergency room visits (0.3% v 0.2%).^15^ The study authors found the relative risk of medical visits or hospitalizations between both groups was the same even when adjusting for age and that age, sex, nor dose had an influence on the severity of influenza like illness symptoms.

### Safety outcomes

Three of the included studies in adult populations reported adverse events that occurred during the trial while one study indicated that no adverse events were recorded for the duration of their trial.^14-17^ All three studies reporting adverse events compared different doses of Fluzone vaccine including 3-μg, 6-μg, 7.5-μg, 9-μg, and 15-μg per strain doses.

Two of the studies were carried out in adult populations and one study was conducted in older adults (>60 years of age).^15-17^ One study found that joint or muscle pain following vaccination was statistically significantly higher in the full dose (15-μg) group compared to the half-dose (7.5-μg) group and that while injection site pain initially appeared to be statistically significantly higher in the full dose group, when adjusted to include only clinically significant pain levels (≥3 out of 5 on a visual analogue scale) the difference was no longer statistically significant.^15^ The study found no differences in occurrence or severity of any other adverse effects. Similarly, one study comparing four different doses of Fluzone (3-μg, 6-μg, 9-μg, and 15-μg per strain) did not report any differences between the IM vaccination groups. .^16^ Finally, the study in older adults also found no difference in the occurrence or severity of adverse events in the low dose (9-μg) versus high dose (15-μg) group and found no serious adverse events that were considered related to the vaccine.^17^

## DISCUSSION

PHAC commissioned this review to identify potentially safe and effective dose-sparing strategies for intramuscular administration of seasonal influenza vaccines in healthy individuals of all ages that have been evaluated in human trials. Thirteen randomized controlled trials published between 2006 and 2019 comparing standard/full-dose and half/low-dose vaccines were included in this scoping review after a comprehensive search of the electronic databases, trial registries and references of relevant systematic reviews. The majority of the included studies were conducted in children and evaluated trivalent influenza vaccines (TIV).

In young children, there were no effectiveness outcomes of interest reported, but local reactogenicity, systemic reactogenicity and adverse events were comparable across the full-dose and half-dose TIV and QIV vaccine arms. In addition, the authors of one study in children and adolescents that compared full-dose QIV using pre-filled syringes (PFS) versus multi-dose vials (MDV) also found no statistically significant differences in safety outcomes between administration formats, suggesting MDV QIV may be a viable alternative format for seasonal influenza vaccination. In adults (including older adults), half-dose QIV was considered equally effective as high-dose in the two studies that assessed clinical effectiveness and safety profiles were similar across groups in all 4 studies.

This rapid scoping review was conducted within a 6-week timeline and the methods were tailored to provide preliminary results to the stakeholders within 4 weeks. We limited the search by date (past 20 years) and language (English), and data charting was conducted by one abstractor and one verifier. In the initial project plan, we outlined that the literature search would be limited to the last 10 years and screening of abstracts and full-texts would be done by a single reviewer, however given the manageable search output we expanded the search to the last 20 years and all screening was completed in duplicate. Also due to the timeline, we limited the number of outcomes of interest. Further exploration of the immunogenicity of the vaccines is warranted, as we did not abstract these results due to the rapid nature of this review. Finally, some dose-sparing studies were not included in the report because they did not include vaccines that were deemed of interest to the stakeholder or the vaccine was unclear. These 12 studies are listed in Appendix C and we have followed-up with the authors of the unclear studies. Given the size of this review, inclusion of these additional studies may impact the results.

## CONCLUSION

Overall there seems to be no significant difference in safety or clinical effectiveness outcomes with the use of low-dose compared to full-dose influenza vaccines, which is promising given the predicted resource constraints in the upcoming influenza season due to the 2019 novel coronavirus (COVID-19). However, due to the low number of studies in adults and the lack of studies assessing confirmed influenza and influenza-like illness, there remains a need for further evaluation of the clinical effectiveness of IM dose-sparing strategies using vaccines currently available in this population. Future research should focus on a systematic review with meta-analysis to confirm the validity of the evidence presented in this rapid scoping review.

## Data Availability

All datasets supporting the conclusions of this article are included within the article.

## Copyright claims/Disclaimers

The intellectual property rights in data and results generated from the work reported in this document are held in joint ownership between the MAGIC team and the named Contributors.

Users are permitted to disseminate the data and results presented in this report provided that the dissemination (i) does not misrepresent the data, results, analyses or conclusions, and (ii) is consistent with academic practice, the rights of any third party publisher, and applicable laws. Any dissemination of the data and results from this document shall properly acknowledge the MAGIC team and named Contributors.

## Acknowledgements

Jessie McGowan (literature search development), Tamara Radar (PRESS of literature search), Alissa Epworth (literature search execution and full-text retrieval), Navjot Mann (author contact and report preparation).

## APPENDIX A MEDLINE search strategy

**Database: Ovid MEDLINE(R) ALL <1946 to May 29, 2020> Search Strategy:**

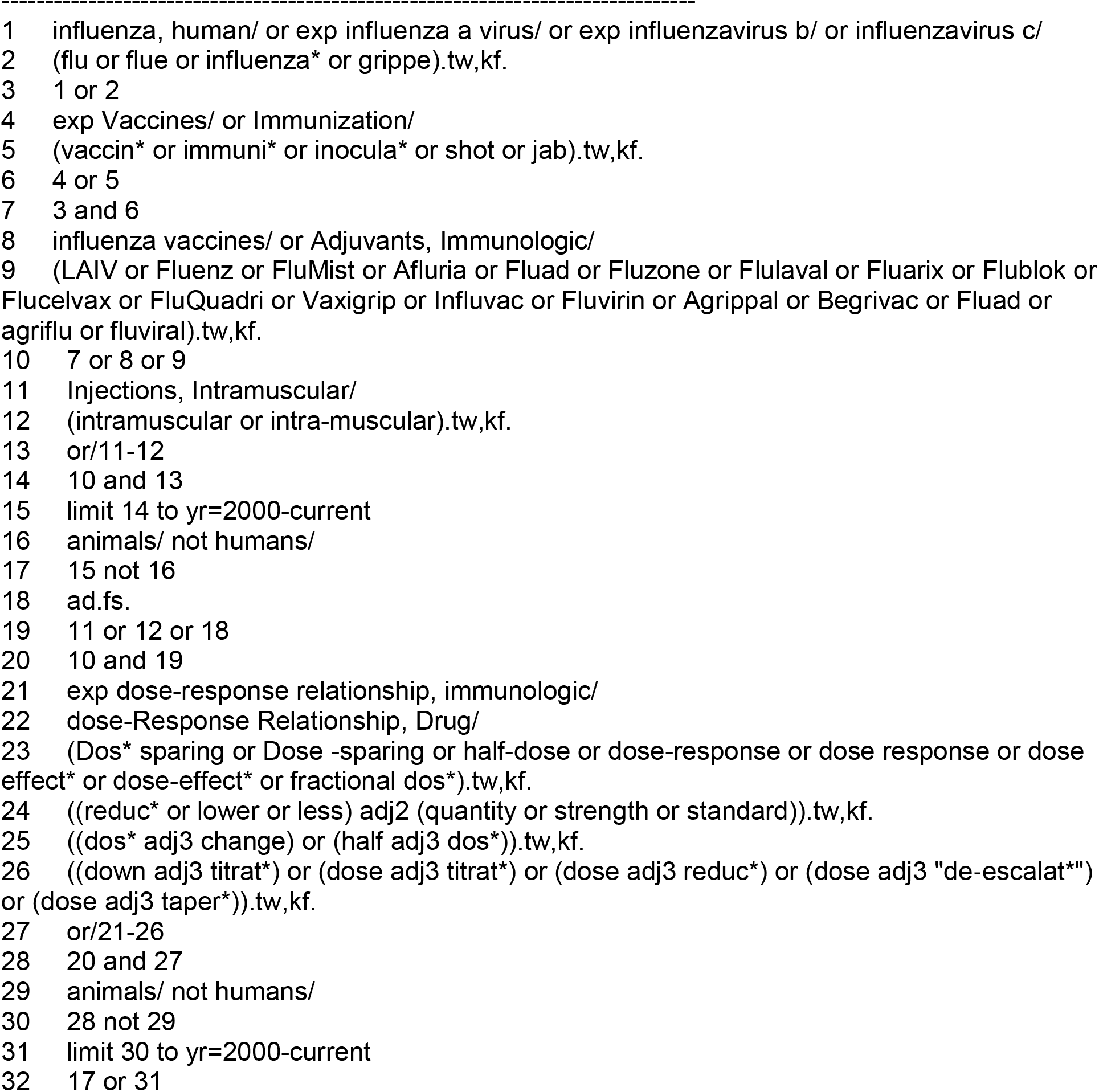

## APPENDIX B List of eligible vaccines

APPENDIX B – List of eligible vaccines
| Product name<br>(manufacturer) | Vaccine Characteristic |  |  |  |  |
| --- | --- | --- | --- | --- | --- |
|  | Vaccine type | Route of administration | Authorized ages for use | Antigen content for each vaccine strain | Formats available |
| Flulaval Tetra (GSK) | IIV4-SD (split virus) | IM | 6 months and older | 15 µg HA /0.5 mL dose | 5 mL multi-dose vial<br>Single dose pre-filled syringe |
| Fluzone Quadrivalent (Sanofi Pasteur) | IIV4-SD (split virus) | IM | 6 months and older | 15 µg HA /0.5 mL dose | 5 mL multi-dose vial<br>Single dose vial<br>Single dose pre-filled syringe without attached needle |
| Afluria Tetra (Seqirus) | IIV4-SD (split virus) | IM | 5 years and older | 15 µg HA /0.5 mL dose | Up to expiry date indicate on vial label |
| Influvac Tetra (BGP Pharma ULC, operating as Mylan) | IIV4-SD (subunit) | IM or deep subcutaneous injection | 3 years and older | 15 µg HA /0.5 mL dose | Single dose pre-filled syringe with or without a needle |
| Vaxigrip Tetra | IIV4 | IM | 6 months and older | Pediatric: 7.5 µg HA /0.25 mL dose<br>Adult: 15 µg HA /0.5 mL dose | 0.5 mL pre-filled syringe |
| Fluarix Tetra/ Influsplit Tetra (GSK) | IIV4 | IM | 6 months and older | 15 µg HA /0.5 mL dose | 0.5 mL pre-filled syringe |
| Agriflu (Seqirus) | IIV3-SD (subunit) | IM | 6 months and older | 15 µg HA /0.5 mL dose | 5 mL multi-dose vial<br>Single dose pre-filled syringe without attached needle |
| Fluad Pediatric and Fluad (Seqirus) | IIV3-Adj (subunit) | IM | Pediatric: 6-23 months<br>Adult: 65 years and older | Pediatric: 7.5 µg HA /0.25 mL dose<br>Adult: 15 µg HA /0.5 mL dose | Single dose pre-filled syringe without a needle |
| Fluviral (GSK) | IIV3-SD (split virus) | IM | 6 months and older | 15 µg HA /0.5 mL dose | 5 mL multi-dose vial |
| Fluzone TIV (Sanofi Pasteur) | IIV3-HD (split virus) | IM | 65 years and older | Adult: 15 µg HA /0.5 mL dose | 0.5 mL pre-filled syringe |
| Vaxigrip TIV | IIV3-SD | IM | 6 months and older | Pediatric: 7.5 µg HA /0.25 mL dose<br>Adult: 15 µg HA /0.5 mL dose | 0.5 mL pre-filled syringe |

**Note:** list of vaccines included in the review is based on feedback from PHAC and the 2020-2021 seasonal vaccine availability in Canada found here: https://www.canada.ca/en/public-health/services/publications/vaccines-immunization/canadian-immunization-guide-statement-seasonal-influenza-vaccine-2020-2021.html#appA

## APPENDIX C Excluded dose-sparing studies

APPENDIX C – Excluded dose-sparing studies
| Reference | Reason for exclusion |
| --- | --- |
| Euctr, H. U. A Randomized, Double-blind, Multi-Center Study to Evaluate Safety and Immunogenicity of One Dose of Four FLUVAL AB-like (Trivalent, Whole Virus, Aluminium Phosphate Gel Adjuvanted) Influenza Vaccines Containing 3.5[micro]gHA, 6[micro]gHA, 9[micro]gHA or 1. 2011. Available from: http://www.who.int/trialsearch/Trial2.aspx?TrialID=EUCTR2011 | exclude - dose-sparing but vaccine not of interest |
| Vajo Z, Tamas F, Jankovics I. A reduced-dose seasonal trivalent influenza vaccine is safe and immunogenic in adult and elderly patients in a randomized controlled trial. *Clin Vaccine Immunol.* 2012;19(3):313-318. doi:10.1128/CVI.05619-11 | exclude - dose-sparing but vaccine not of interest |
| Treanor J, Keitel W, Belshe R, et al. Evaluation of a single dose of half strength inactivated influenza vaccine in healthy adults. *Vaccine.* 2002;20(7-8):1099-1105. doi:10.1016/s0264-410x(01)00440-6 | exclude - dose-sparing but vaccine not of interest |
| Euctr. A Randomized, Active Controlled, Double-blind, Multi-Centre Study to Evaluate Safety and Immunogenicity of One Dose of FLUVAL AB-like (Trivalent, Whole Virus, Aluminium Phosphate Gel Adjuvanted) Influenza Vaccine Containing 6µgHA of Seasonal A/H1N1, A/H3N2 and B Influenza Antigens in Non-elderly Adult and Elderly Subjects. 2011. Available from: http://www.who.int/trialsearch/Trial2.aspx?TrialID=EUCTR2011-003314-16-HU | exclude - dose-sparing but experimental vaccine |
| Euctr, E. S. Clinical study to compare the safety of two influenza vaccines in children and adolescents of 3 to less than 18 years of age at risk for influenza-related complications. 2013. Available from: http://www.who.int/trialsearch/Trial2.aspx?TrialID=EUCTR2013 | exclude - dose-sparing but experimental vaccine |
| Pillet S, Aubin É, Trépanier S, et al. A plant-derived quadrivalent virus like particle influenza vaccine induces cross-reactive antibody and T cell response in healthy adults. *Clin Immunol.* 2016;168:72-87. doi:10.1016/j.clim.2016.03.008 | exclude - dose-sparing but experimental vaccine |
| Lee JH, Cho HK, Kim KH, et al. Evaluation of Waning Immunity at 6 Months after Both Trivalent and Quadrivalent Influenza Vaccination in Korean Children Aged 6-35 Months. *J Korean Med Sci.* 2019;34(46):e279. Published 2019 Dec 2. doi:10.3346/jkms.2019.34.e279 | exclude - dose-sparing but experimental vaccine |
| Treanor JJ, Taylor DN, Tussey L, et al. Safety and immunogenicity of a recombinant hemagglutinin influenza-flagellin fusion vaccine (VAX125) in healthy young adults. *Vaccine.* 2010;28(52):8268-8274. doi:10.1016/j.vaccine.2010.10.009 | exclude - dose-sparing but experimental vaccine |
| Vajo Z, Balaton G, Vajo P, Kalabay L, Erdman A, Torzsa P. Dose sparing and the lack of a dose-response relationship with an influenza vaccine in adult and elderly patients - a randomized, double-blind clinical trial. *Br J Clin Pharmacol.* 2017;83(9):1912-1920. doi:10.1111/bcp.13289 | exclude - dose-sparing but vaccine not of interest |
| Ctri. Study of a Single Dose or Two Doses of a Quadrivalent Influenza Vaccine in Subjects Aged 6 Months or Older in India. 2015. Available from: http://www.who.int/trialsearch/Trial2.aspx?TrialID=CTRI | exclude - dose-sparing but unclear vaccine (waiting for author response) |
| Euctr, F. I. Safety and Immunogenicity of the Quadrivalent Influenza Vaccine Administered via the Intramuscular Route in Children Aged 3 to 8 Years. 2011. Available from: http://www.who.int/trialsearch/Trial2.aspx?TrialID=EUCTR2011 | exclude - dose-sparing but unclear vaccine (waiting for author response) |
| Euctr, C. Z. A randomized, double-blind, placebo-controlled, multi-country and multi-center, phase IV study to demonstrate the efficacy of GSK Biologicals' influenza vaccine (Fluarix[TM]) administered intramuscularly in adults. - FluarixUS-006. 2006. Available from: http://www.who.int/trialsearch/Trial2.aspx?TrialID=EUCTR2006 | exclude - dose-sparing but unclear vaccine (waiting for author response) |

## APPENDIX D Study and patient data

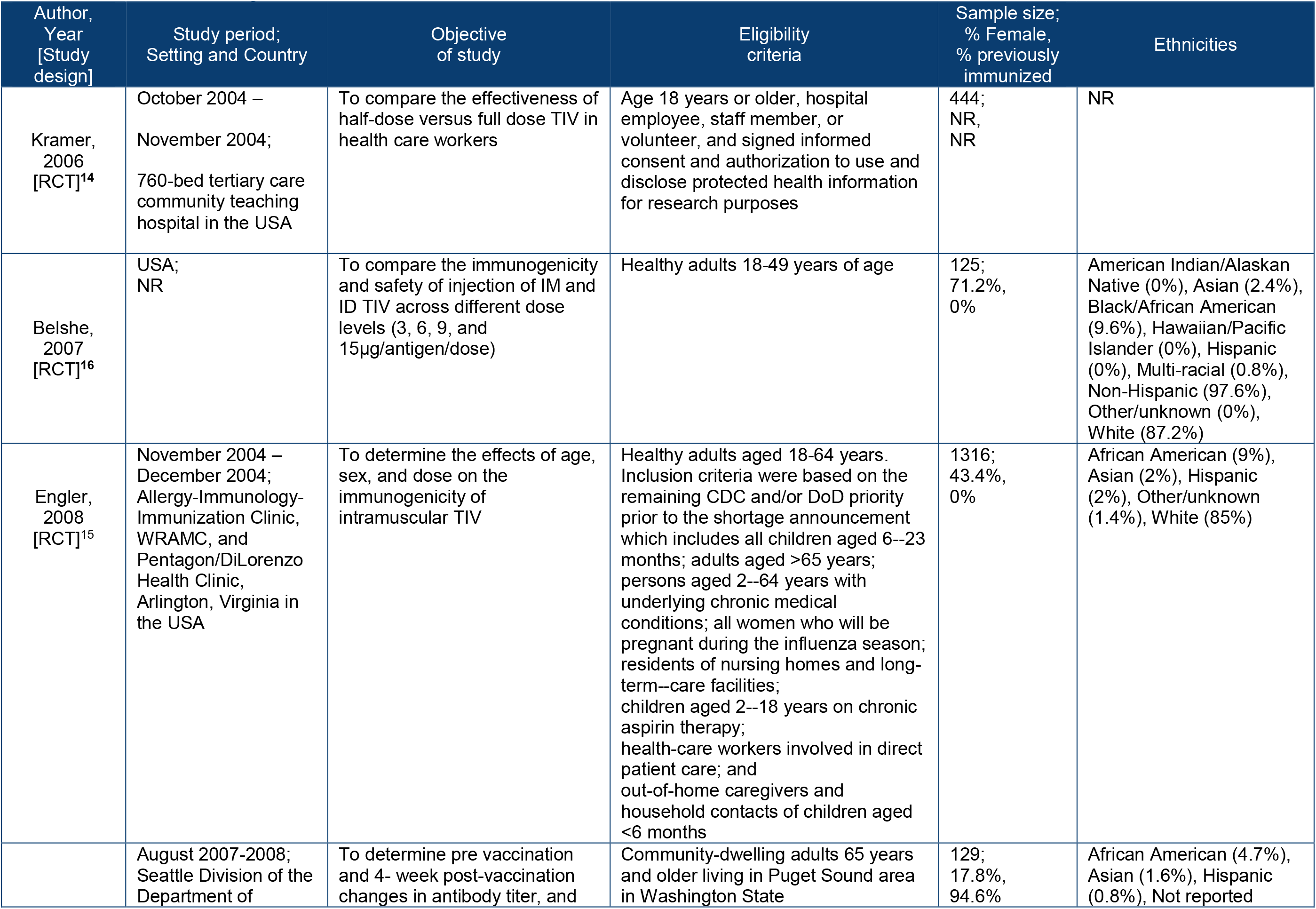
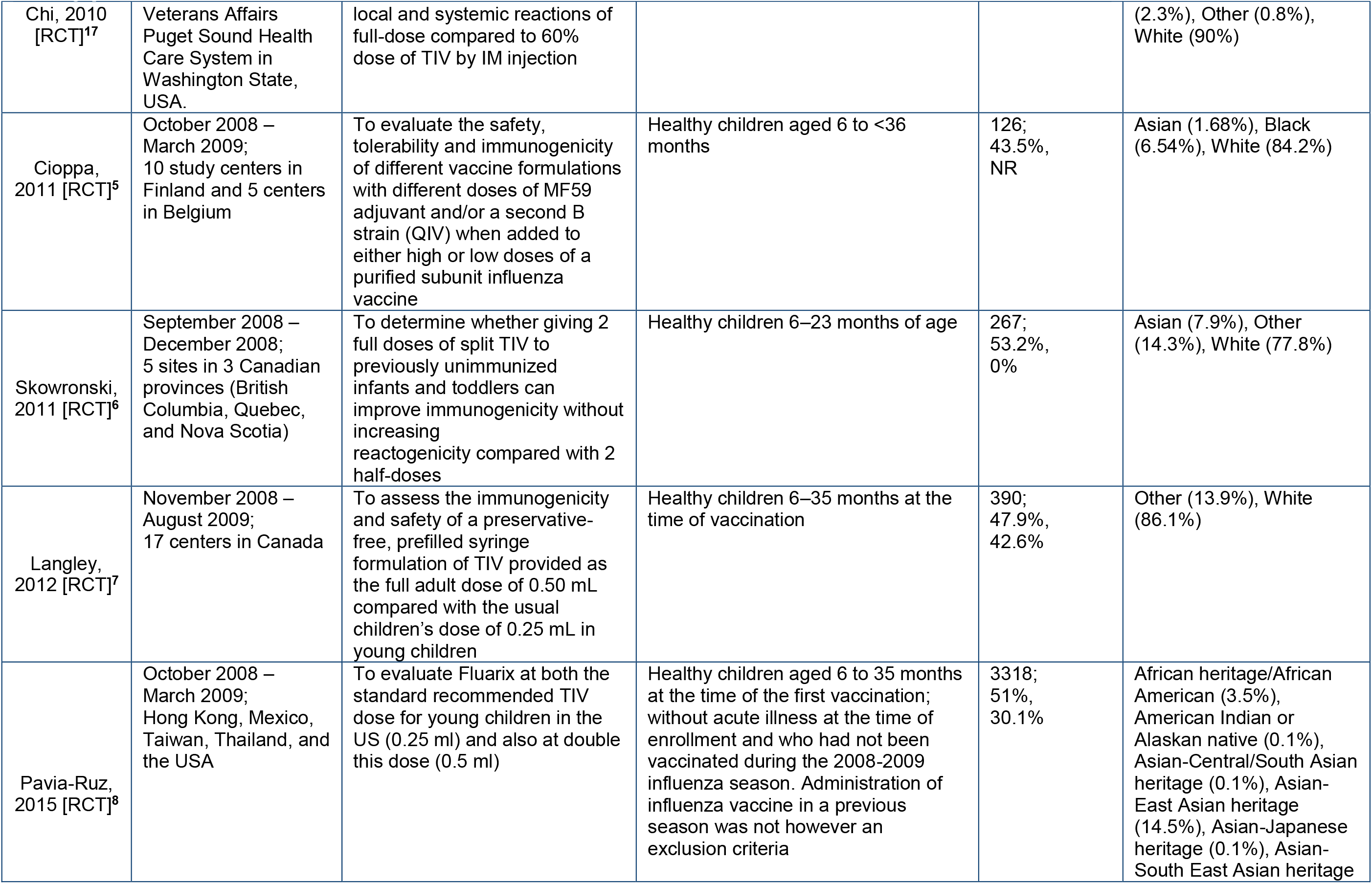
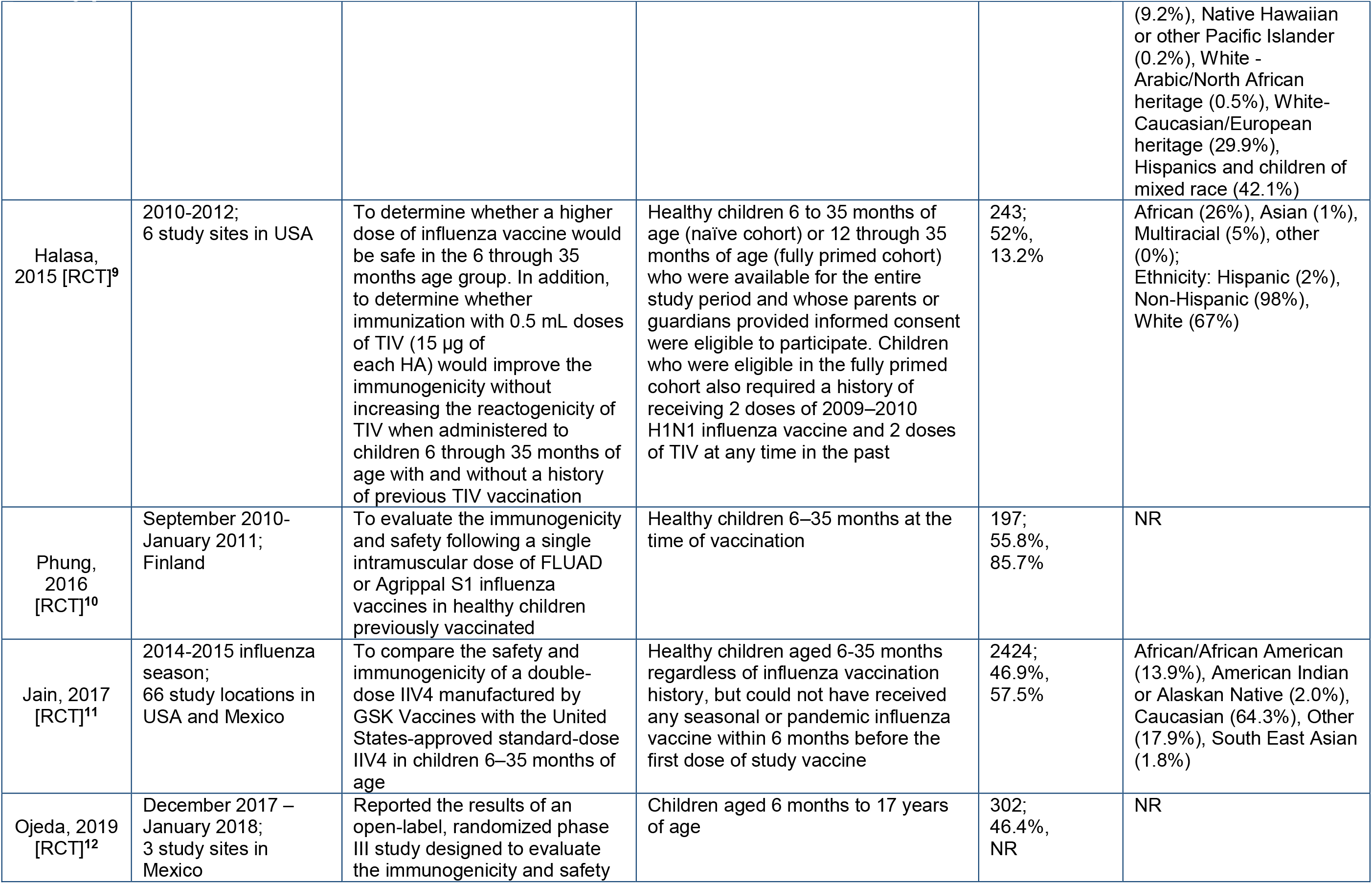
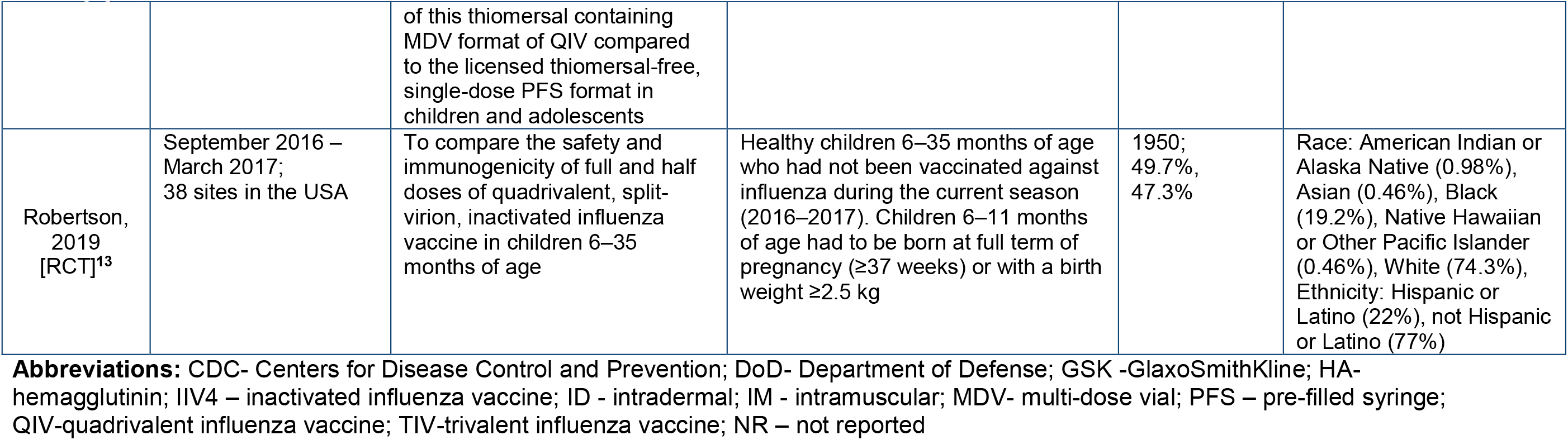

APPENDIX D – Study and patient data
| Author, Year [Study design] | Study period; Setting and Country | Objective of study | Eligibility criteria | Sample size; % Female, % previously immunized | Ethnicities |
| --- | --- | --- | --- | --- | --- |
| Kramer, 2006 [RCT]<sup>14</sup> | October 2004 – November 2004;<br>760-bed tertiary care community teaching hospital in the USA | To compare the effectiveness of half-dose versus full dose TIV in health care workers | Age 18 years or older, hospital employee, staff member, or volunteer, and signed informed consent and authorization to use and disclose protected health information for research purposes | 444;<br>NR,<br>NR | NR |
| Belshe, 2007 [RCT]<sup>16</sup> | USA;<br>NR | To compare the immunogenicity and safety of injection of IM and ID TIV across different dose levels (3, 6, 9, and 15µg/antigen/dose) | Healthy adults 18-49 years of age | 125;<br>71.2%,<br>0% | American Indian/Alaskan Native (0%), Asian (2.4%), Black/African American (9.6%), Hawaiian/Pacific Islander (0%), Hispanic (0%), Multi-racial (0.8%), Non-Hispanic (97.6%), Other/unknown (0%), White (87.2%) |
| Engler, 2008 [RCT]<sup>15</sup> | November 2004 – December 2004;<br>Allergy-Immunology-Immunization Clinic, WRAMC, and Pentagon/DiLorenzo Health Clinic, Arlington, Virginia in the USA | To determine the effects of age, sex, and dose on the immunogenicity of intramuscular TIV | Healthy adults aged 18-64 years. Inclusion criteria were based on the remaining CDC and/or DoD priority prior to the shortage announcement which includes all children aged 6--23 months; adults aged >65 years; persons aged 2--64 years with underlying chronic medical conditions; all women who will be pregnant during the influenza season; residents of nursing homes and long-term--care facilities; children aged 2--18 years on chronic aspirin therapy; health-care workers involved in direct patient care; and out-of-home caregivers and household contacts of children aged <6 months | 1316;<br>43.4%,<br>0% | African American (9%), Asian (2%), Hispanic (2%), Other/unknown (1.4%), White (85%) |
|  | August 2007-2008;<br>Seattle Division of the Department of | To determine pre vaccination and 4- week post-vaccination changes in antibody titer, and | Community-dwelling adults 65 years and older living in Puget Sound area in Washington State | 129;<br>17.8%,<br>94.6% | African American (4.7%), Asian (1.6%), Hispanic (0.8%), Not reported |

| Author,<br>Year<br>[Study<br>design] | Study period;<br>Setting and Country | Objective<br>of study | Eligibility<br>criteria | Sample size;<br>% Female,<br>% previously<br>immunized | Ethnicities |
| --- | --- | --- | --- | --- | --- |
| Chi, 2010<br>[RCT]<sup>17</sup> | Veterans Affairs<br>Puget Sound Health<br>Care System in<br>Washington State,<br>USA. | local and systemic reactions of<br>full-dose compared to 60%<br>dose of TIV by IM injection |  |  | (2.3%), Other (0.8%),<br>White (90%) |
| Cioppa,<br>2011 [RCT]<sup>5</sup> | October 2008 –<br>March 2009;<br>10 study centers in<br>Finland and 5 centers<br>in Belgium | To evaluate the safety,<br>tolerability and immunogenicity<br>of different vaccine formulations<br>with different doses of MF59<br>adjuvant and/or a second B<br>strain (QIV) when added to<br>either high or low doses of a<br>purified subunit influenza<br>vaccine | Healthy children aged 6 to <36<br>months | 126;<br>43.5%,<br>NR | Asian (1.68%), Black<br>(6.54%), White (84.2%) |
| Skowronski,<br>2011 [RCT]<sup>6</sup> | September 2008 –<br>December 2008;<br>5 sites in 3 Canadian<br>provinces (British<br>Columbia, Quebec,<br>and Nova Scotia) | To determine whether giving 2<br>full doses of split TIV to<br>previously unimmunized<br>infants and toddlers can<br>improve immunogenicity without<br>increasing<br>reactogenicity compared with 2<br>half-doses | Healthy children 6–23 months of age | 267;<br>53.2%,<br>0% | Asian (7.9%), Other<br>(14.3%), White (77.8%) |
| Langley,<br>2012 [RCT]<sup>7</sup> | November 2008 –<br>August 2009;<br>17 centers in Canada | To assess the immunogenicity<br>and safety of a preservative-<br>free, prefilled syringe<br>formulation of TIV provided as<br>the full adult dose of 0.50 mL<br>compared with the usual<br>children's dose of 0.25 mL in<br>young children | Healthy children 6–35 months at the<br>time of vaccination | 390;<br>47.9%,<br>42.6% | Other (13.9%), White<br>(86.1%) |
| Pavia-Ruz,<br>2015 [RCT]<sup>8</sup> | October 2008 –<br>March 2009;<br>Hong Kong, Mexico,<br>Taiwan, Thailand, and<br>the USA | To evaluate Fluarix at both the<br>standard recommended TIV<br>dose for young children in the<br>US (0.25 ml) and also at double<br>this dose (0.5 ml) | Healthy children aged 6 to 35 months<br>at the time of the first vaccination;<br>without acute illness at the time of<br>enrollment and who had not been<br>vaccinated during the 2008-2009<br>influenza season. Administration of<br>influenza vaccine in a previous<br>season was not however an<br>exclusion criteria | 3318;<br>51%,<br>30.1% | African heritage/African<br>American (3.5%),<br>American Indian or<br>Alaskan native (0.1%),<br>Asian-Central/South Asian<br>heritage (0.1%), Asian-East Asian heritage<br>(14.5%), Asian-Japanese<br>heritage (0.1%), Asian-South East Asian heritage |
|  |  |  |  |  | (9.2%), Native Hawaiian or other Pacific Islander (0.2%), White - Arabic/North African heritage (0.5%), White-Caucasian/European heritage (29.9%), Hispanics and children of mixed race (42.1%) |
| Halasa, 2015 [RCT]<sup>9</sup> | 2010-2012; 6 study sites in USA | To determine whether a higher dose of influenza vaccine would be safe in the 6 through 35 months age group. In addition, to determine whether immunization with 0.5 mL doses of TIV (15 µg of each HA) would improve the immunogenicity without increasing the reactogenicity of TIV when administered to children 6 through 35 months of age with and without a history of previous TIV vaccination | Healthy children 6 to 35 months of age (naïve cohort) or 12 through 35 months of age (fully primed cohort) who were available for the entire study period and whose parents or guardians provided informed consent were eligible to participate. Children who were eligible in the fully primed cohort also required a history of receiving 2 doses of 2009–2010 H1N1 influenza vaccine and 2 doses of TIV at any time in the past | 243; 52%, 13.2% | African (26%), Asian (1%), Multiracial (5%), other (0%); Ethnicity: Hispanic (2%), Non-Hispanic (98%), White (67%) |
| Phung, 2016 [RCT]<sup>10</sup> | September 2010-January 2011; Finland | To evaluate the immunogenicity and safety following a single intramuscular dose of FLUAD or Agrippal S1 influenza vaccines in healthy children previously vaccinated | Healthy children 6–35 months at the time of vaccination | 197; 55.8%, 85.7% | NR |
| Jain, 2017 [RCT]<sup>11</sup> | 2014-2015 influenza season; 66 study locations in USA and Mexico | To compare the safety and immunogenicity of a double-dose IIV4 manufactured by GSK Vaccines with the United States-approved standard-dose IIV4 in children 6–35 months of age | Healthy children aged 6-35 months regardless of influenza vaccination history, but could not have received any seasonal or pandemic influenza vaccine within 6 months before the first dose of study vaccine | 2424; 46.9%, 57.5% | African/African American (13.9%), American Indian or Alaskan Native (2.0%), Caucasian (64.3%), Other (17.9%), South East Asian (1.8%) |
| Ojeda, 2019 [RCT]<sup>12</sup> | December 2017 – January 2018; 3 study sites in Mexico | Reported the results of an open-label, randomized phase III study designed to evaluate the immunogenicity and safety | Children aged 6 months to 17 years of age | 302; 46.4%, NR | NR |

| Author,<br>Year<br>[Study<br>design] | Study period;<br>Setting and Country | Objective<br>of study | Eligibility<br>criteria | Sample size;<br>% Female,<br>% previously<br>immunized | Ethnicities |
| --- | --- | --- | --- | --- | --- |
|  |  | of this thiomersal containing MDV format of QIV compared to the licensed thiomersal-free, single-dose PFS format in children and adolescents |  |  |  |
| Robertson,<br>2019<br>[RCT]<sup>13</sup> | September 2016 –<br>March 2017;<br>38 sites in the USA | To compare the safety and immunogenicity of full and half doses of quadrivalent, split-virion, inactivated influenza vaccine in children 6–35 months of age | Healthy children 6–35 months of age who had not been vaccinated against influenza during the current season (2016–2017). Children 6–11 months of age had to be born at full term of pregnancy (≥37 weeks) or with a birth weight ≥2.5 kg | 1950;<br>49.7%,<br>47.3% | Race: American Indian or Alaska Native (0.98%), Asian (0.46%), Black (19.2%), Native Hawaiian or Other Pacific Islander (0.46%), White (74.3%), Ethnicity: Hispanic or Latino (22%), not Hispanic or Latino (77%) |

**Abbreviations:** CDC- Centers for Disease Control and Prevention; DoD- Department of Defense; GSK -GlaxoSmithKline; HA- hemagglutinin; IIV4 – inactivated influenza vaccine; ID - intradermal; IM - intramuscular; MDV- multi-dose vial; PFS – pre-filled syringe; QIV-quadrivalent influenza vaccine; TIV-trivalent influenza vaccine; NR – not reported

## APPENDIX E Treatment and outcome data

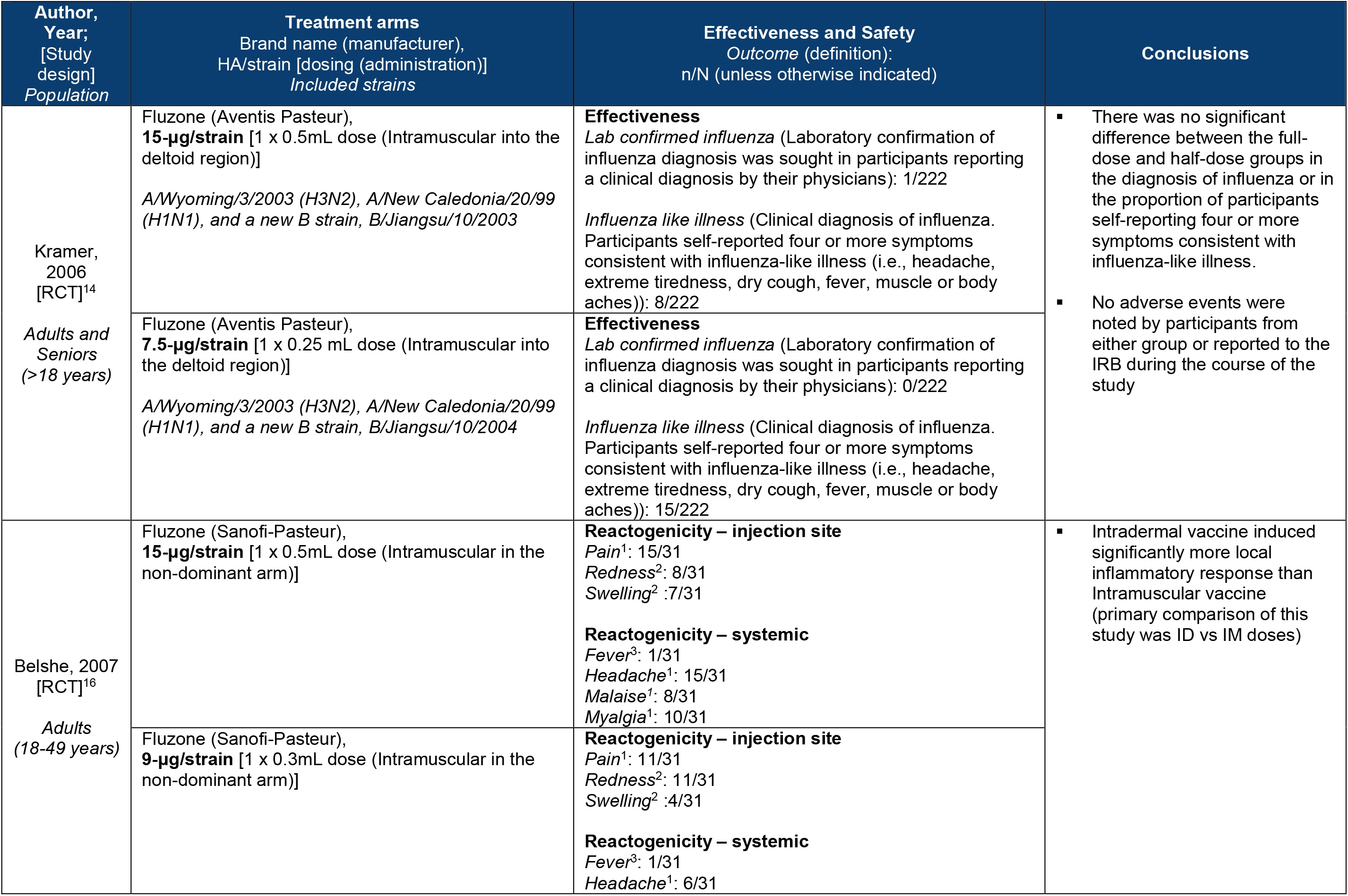
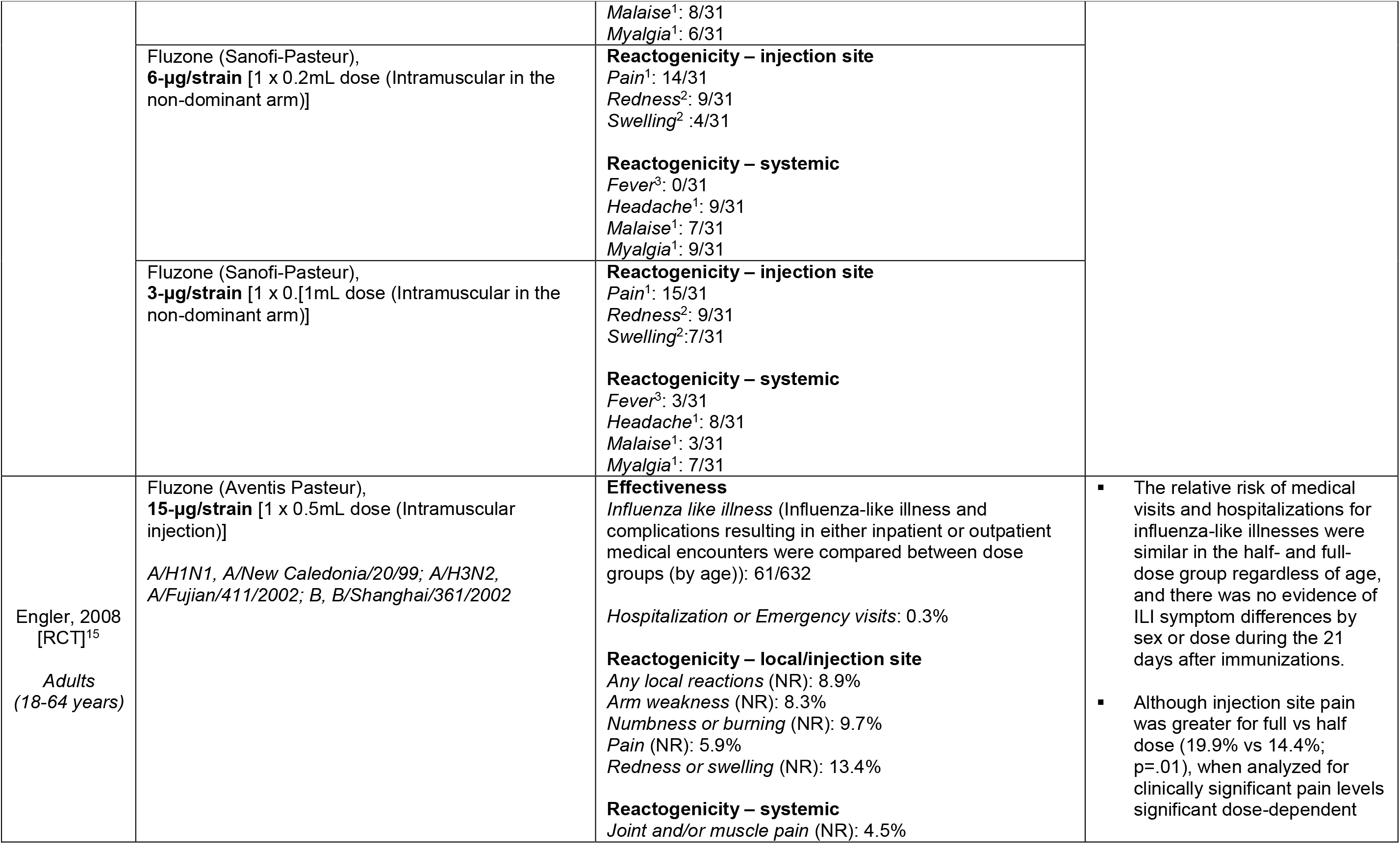
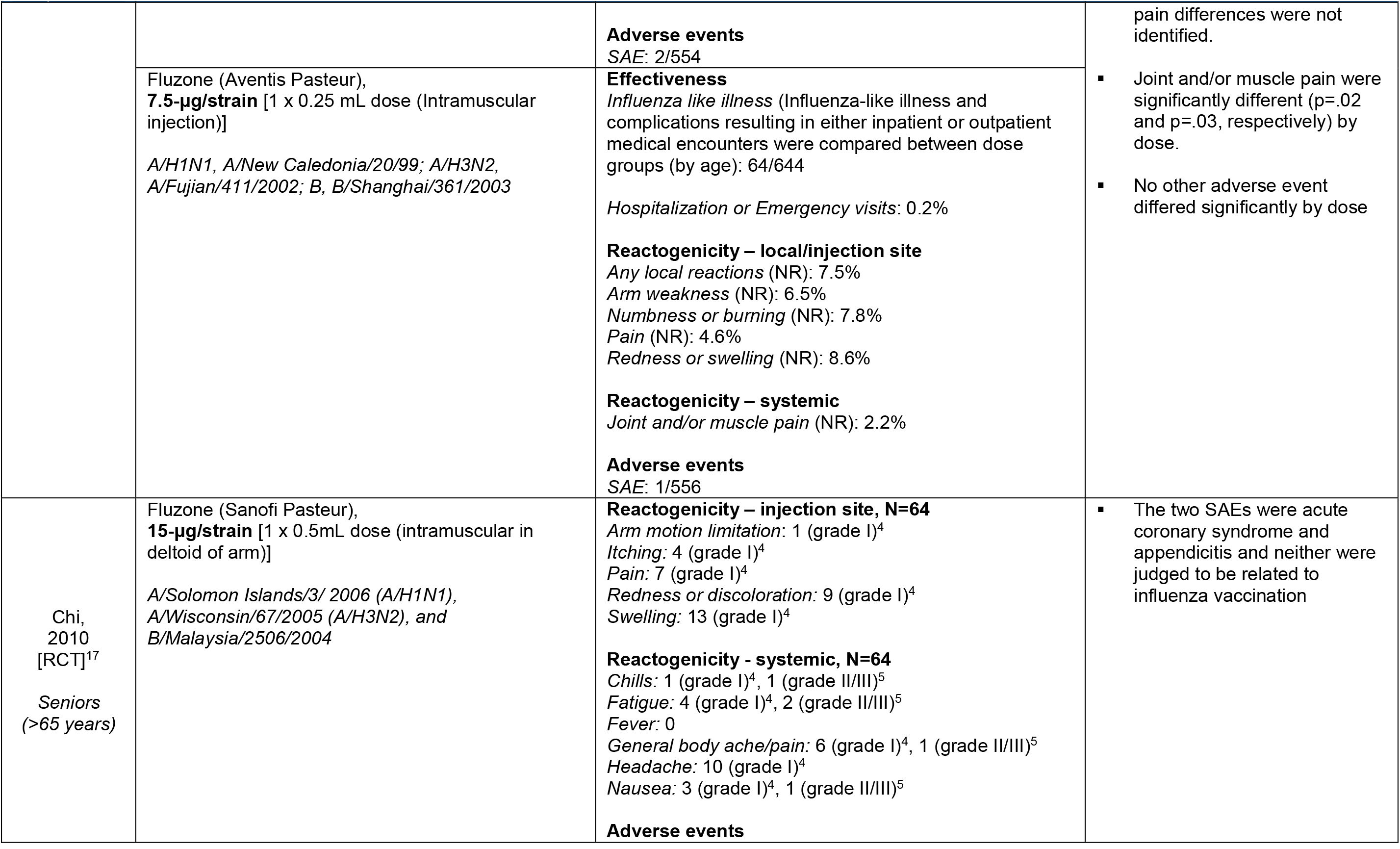
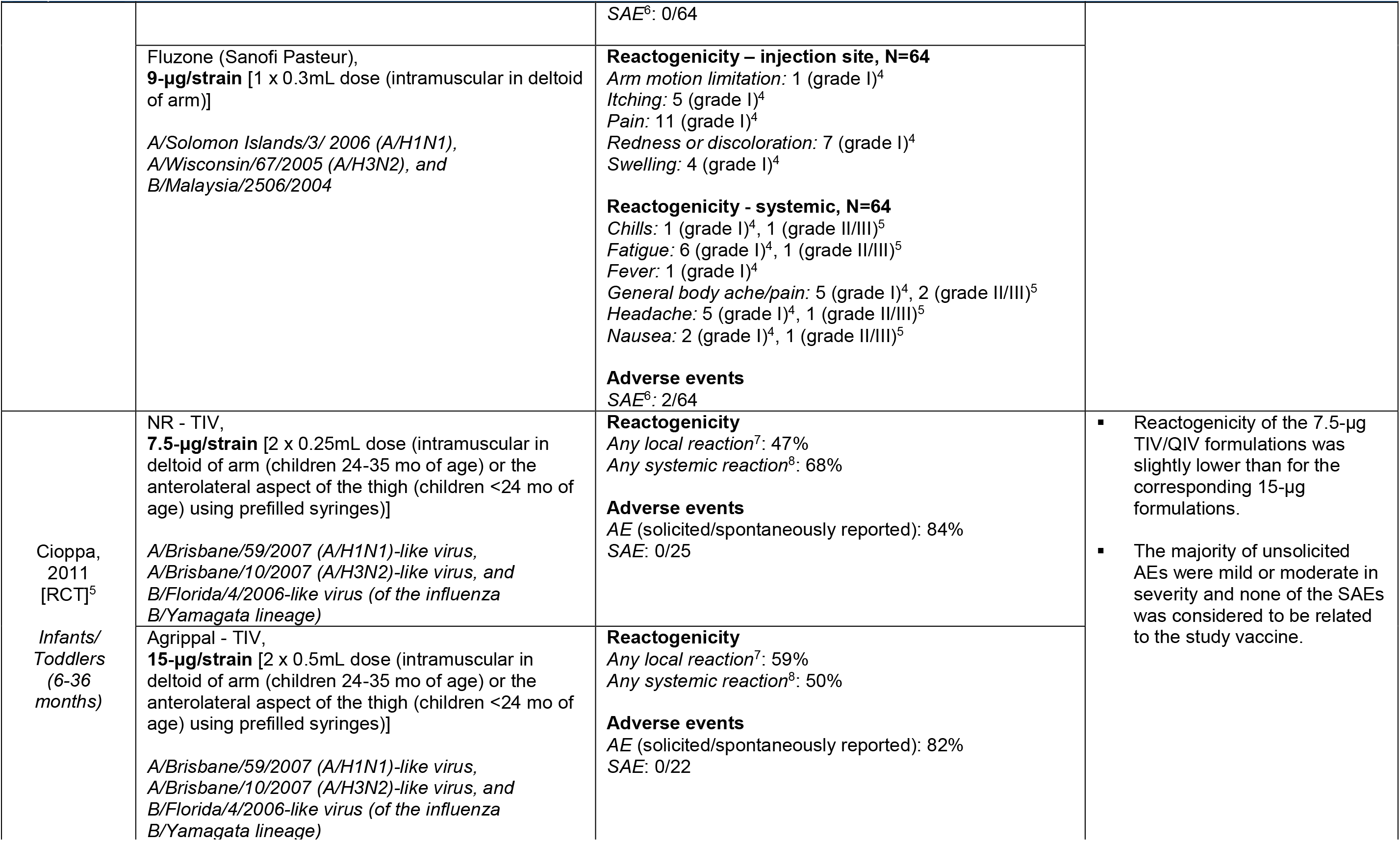
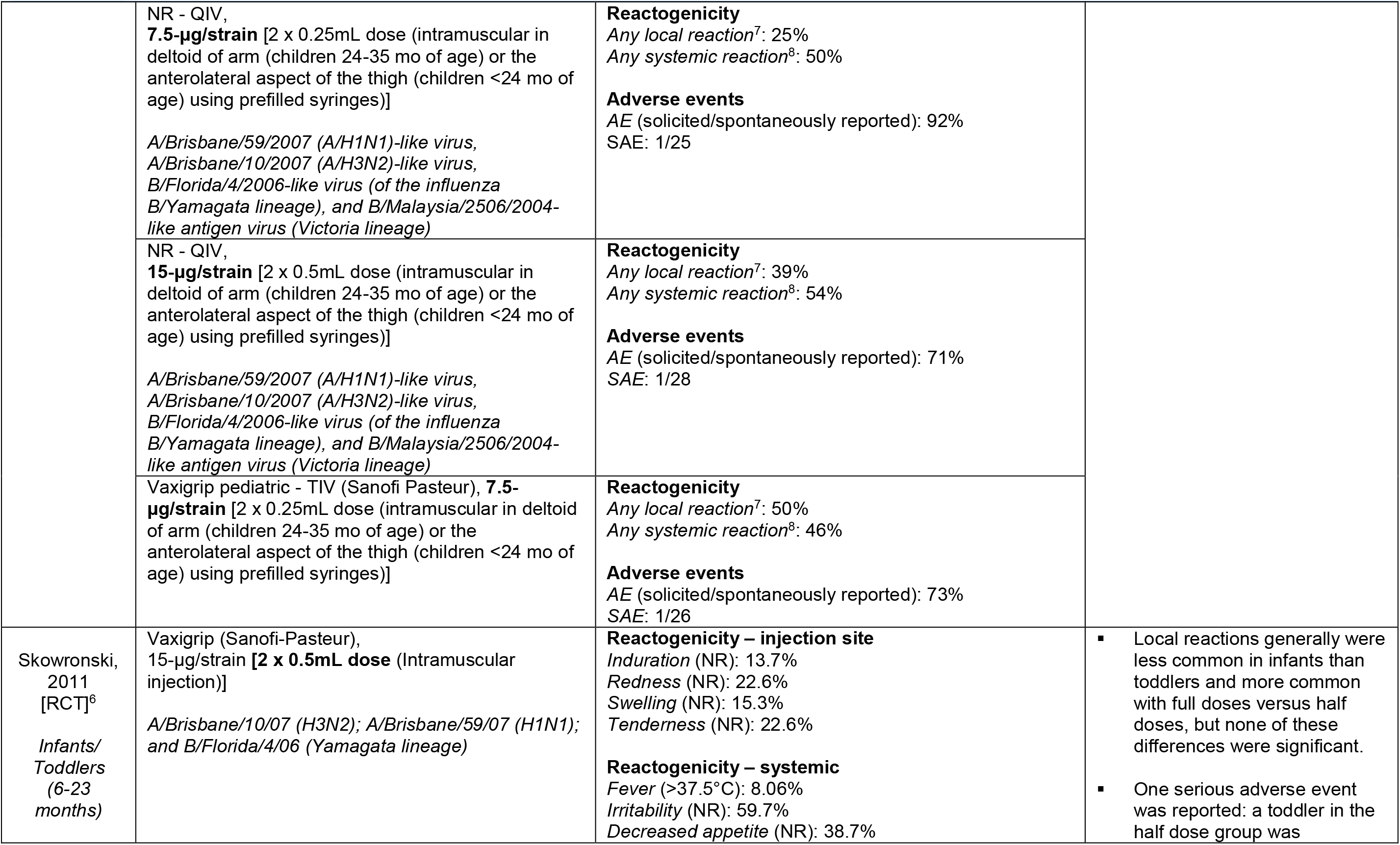
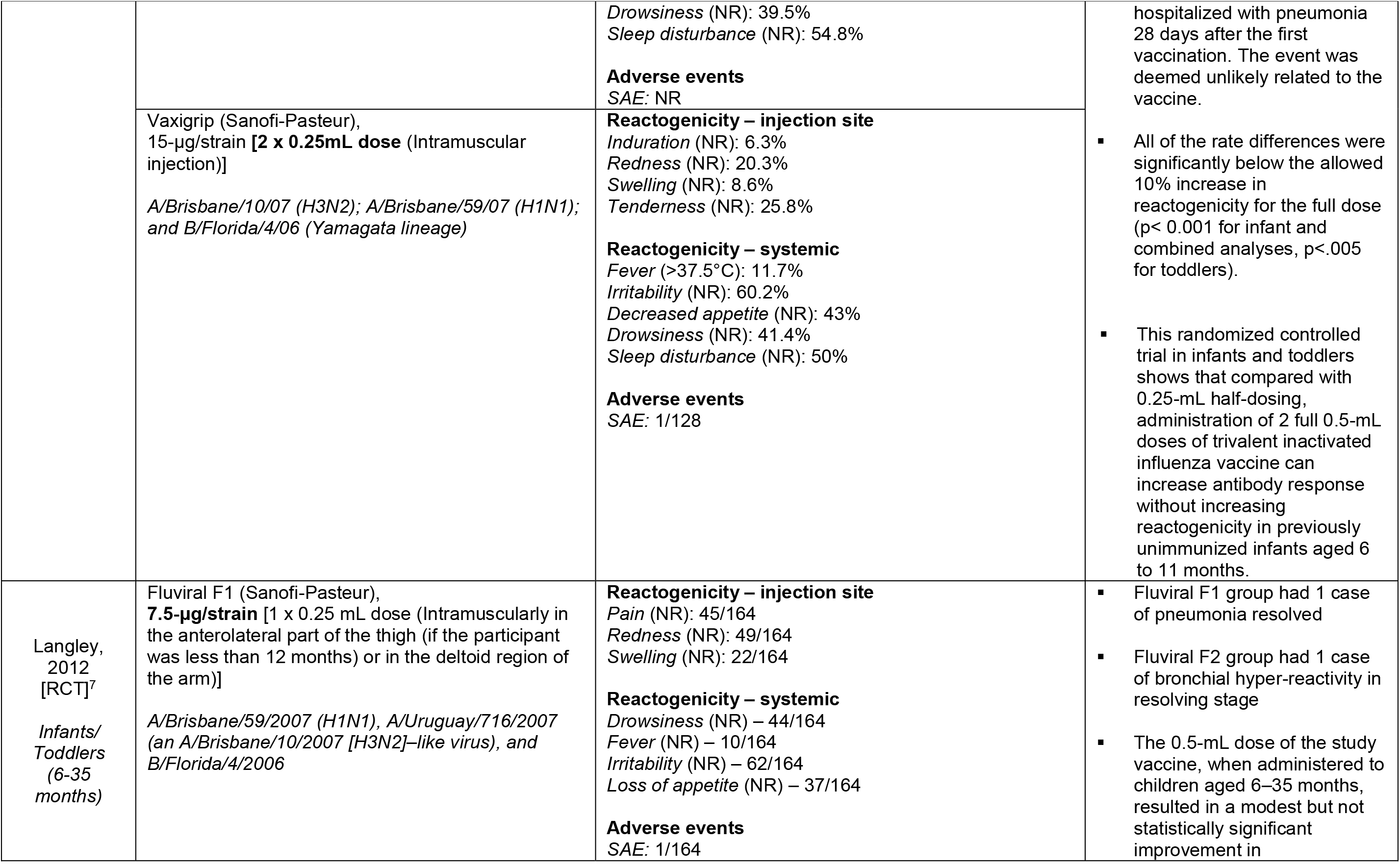
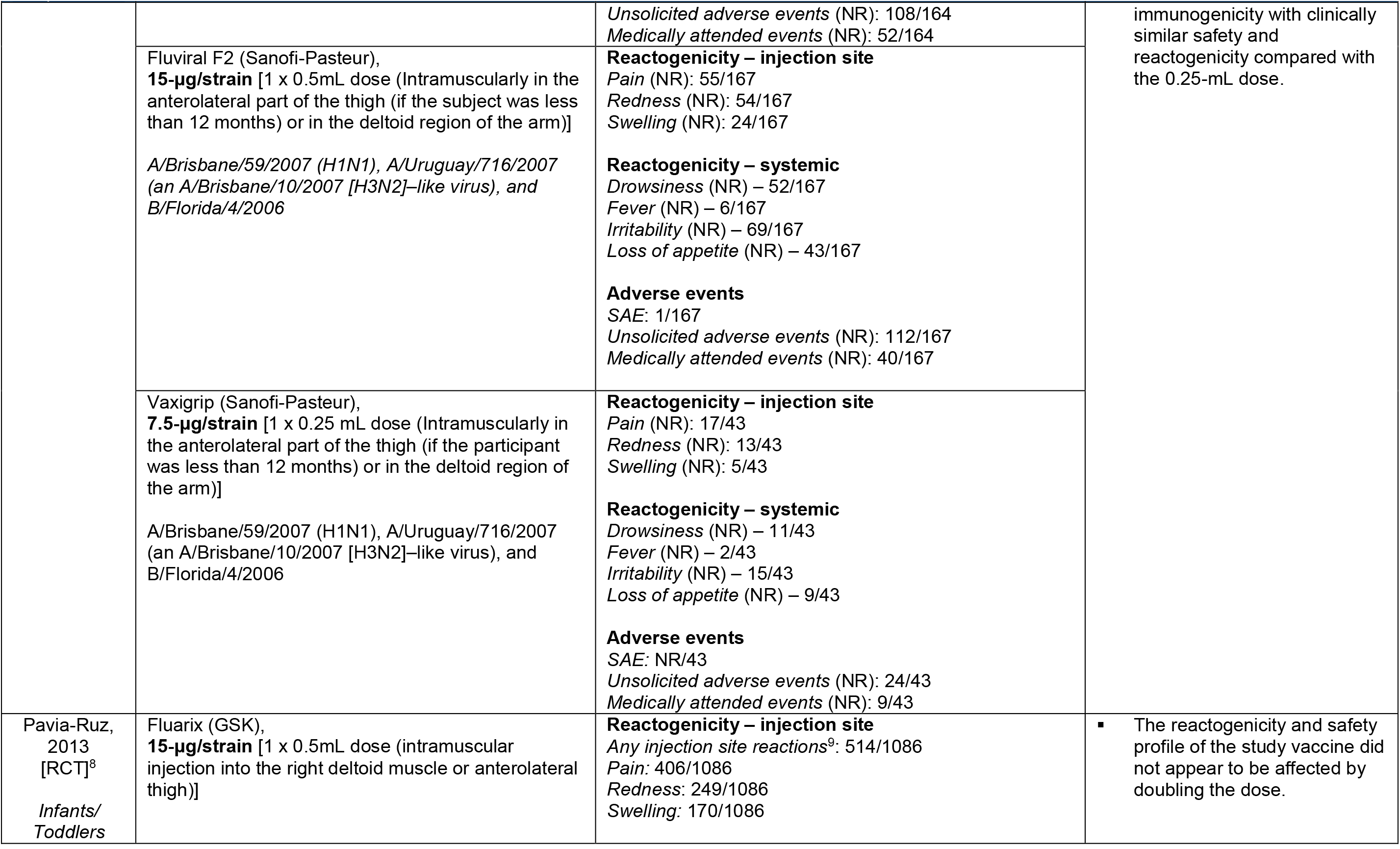
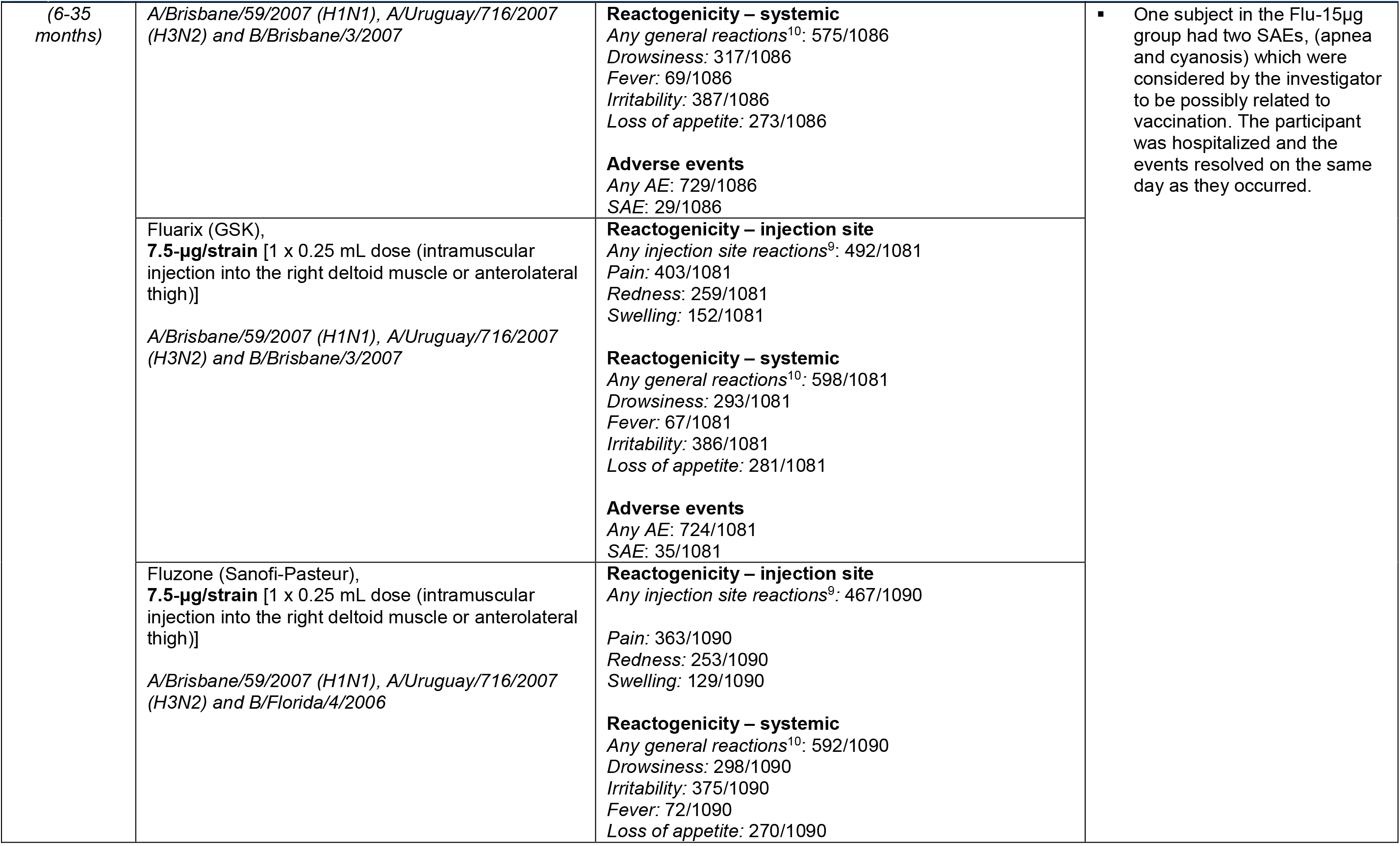
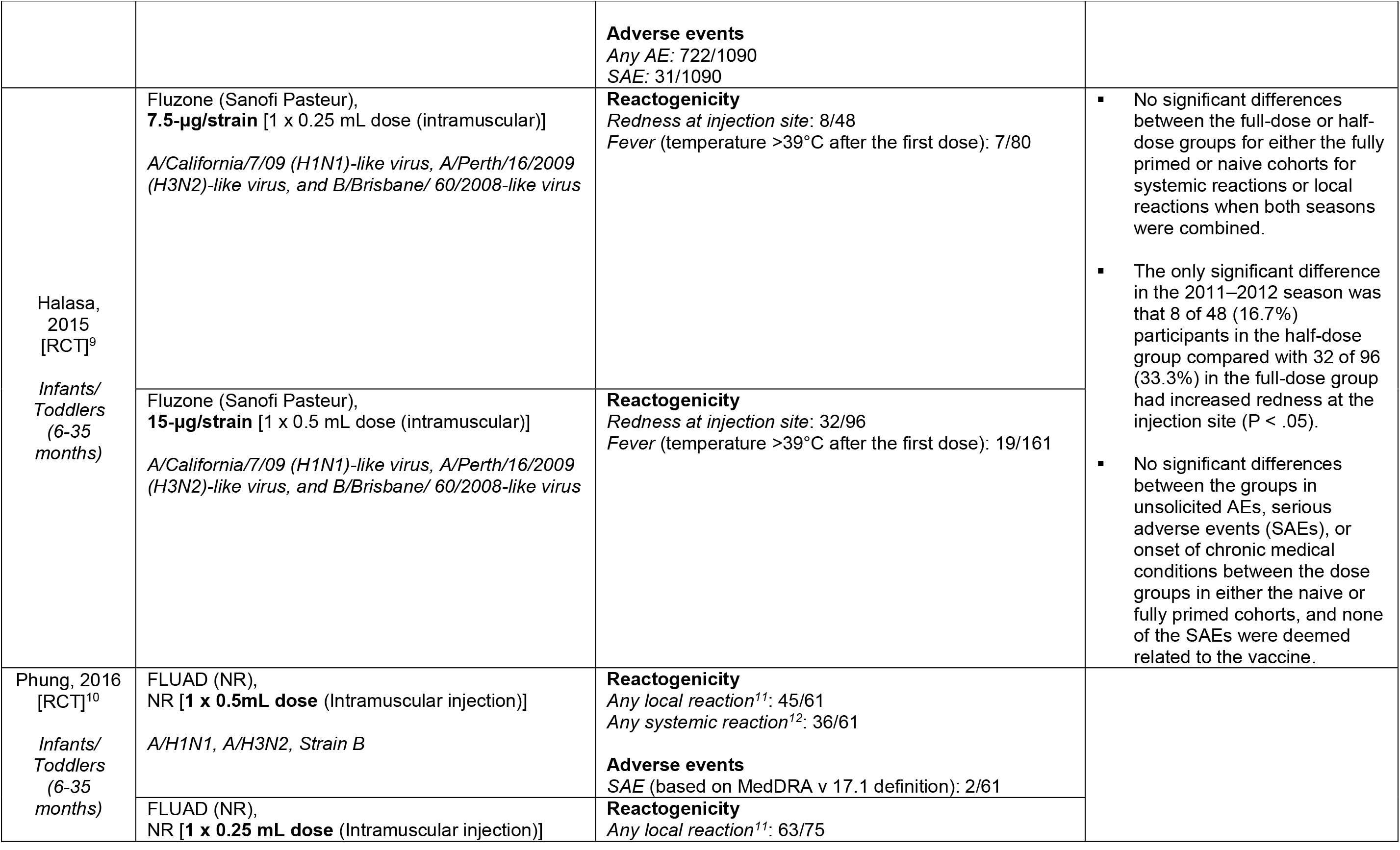
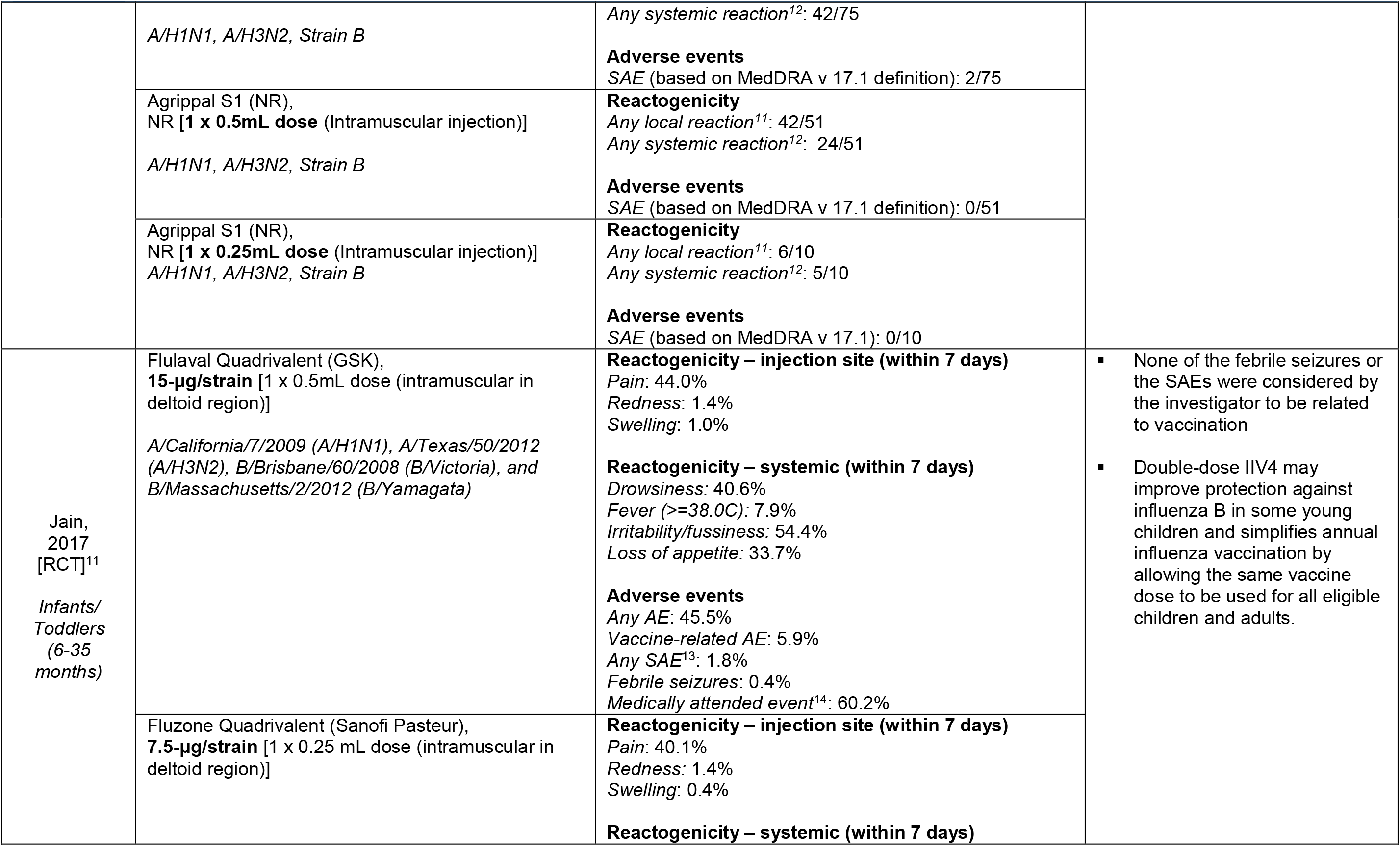
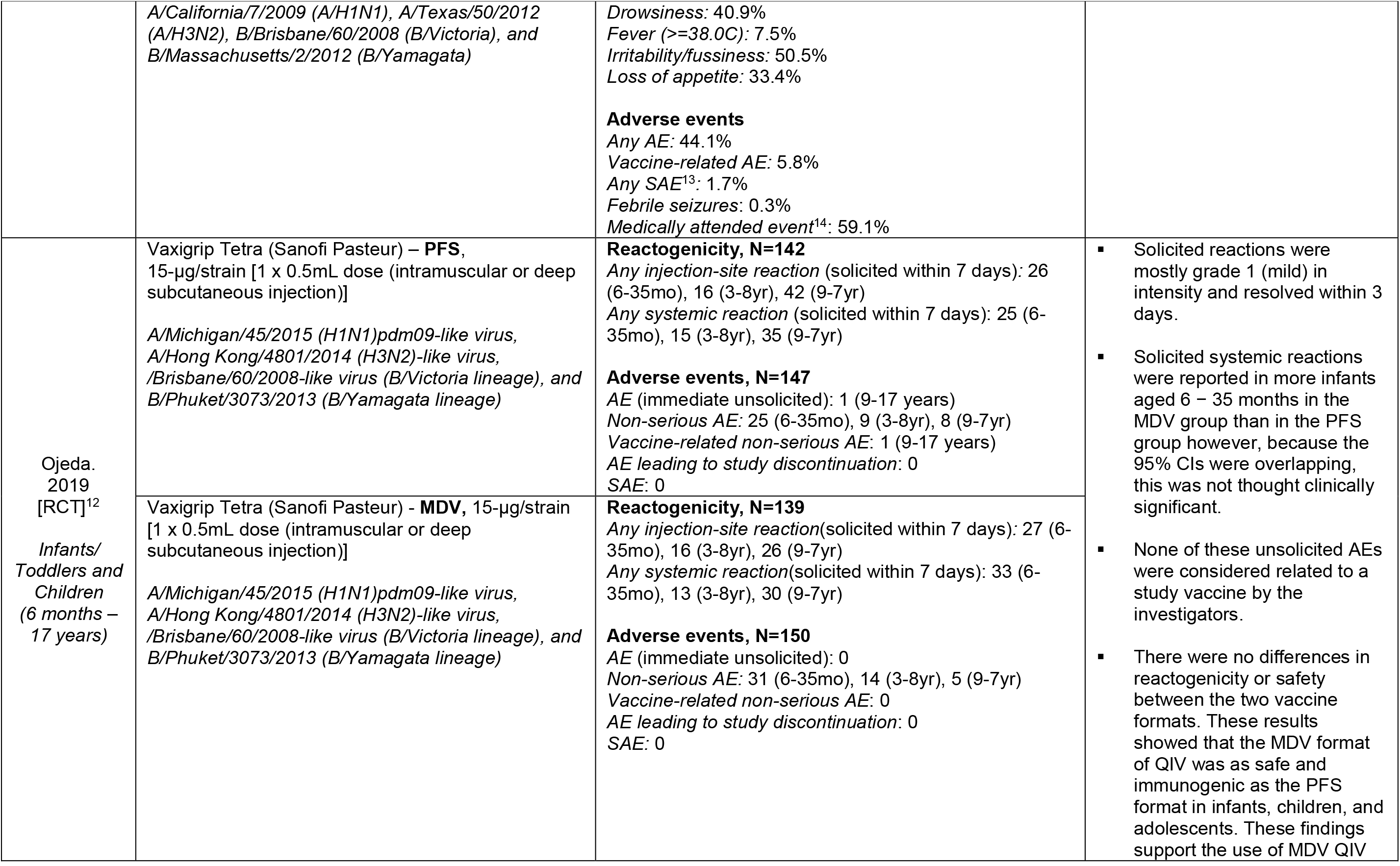
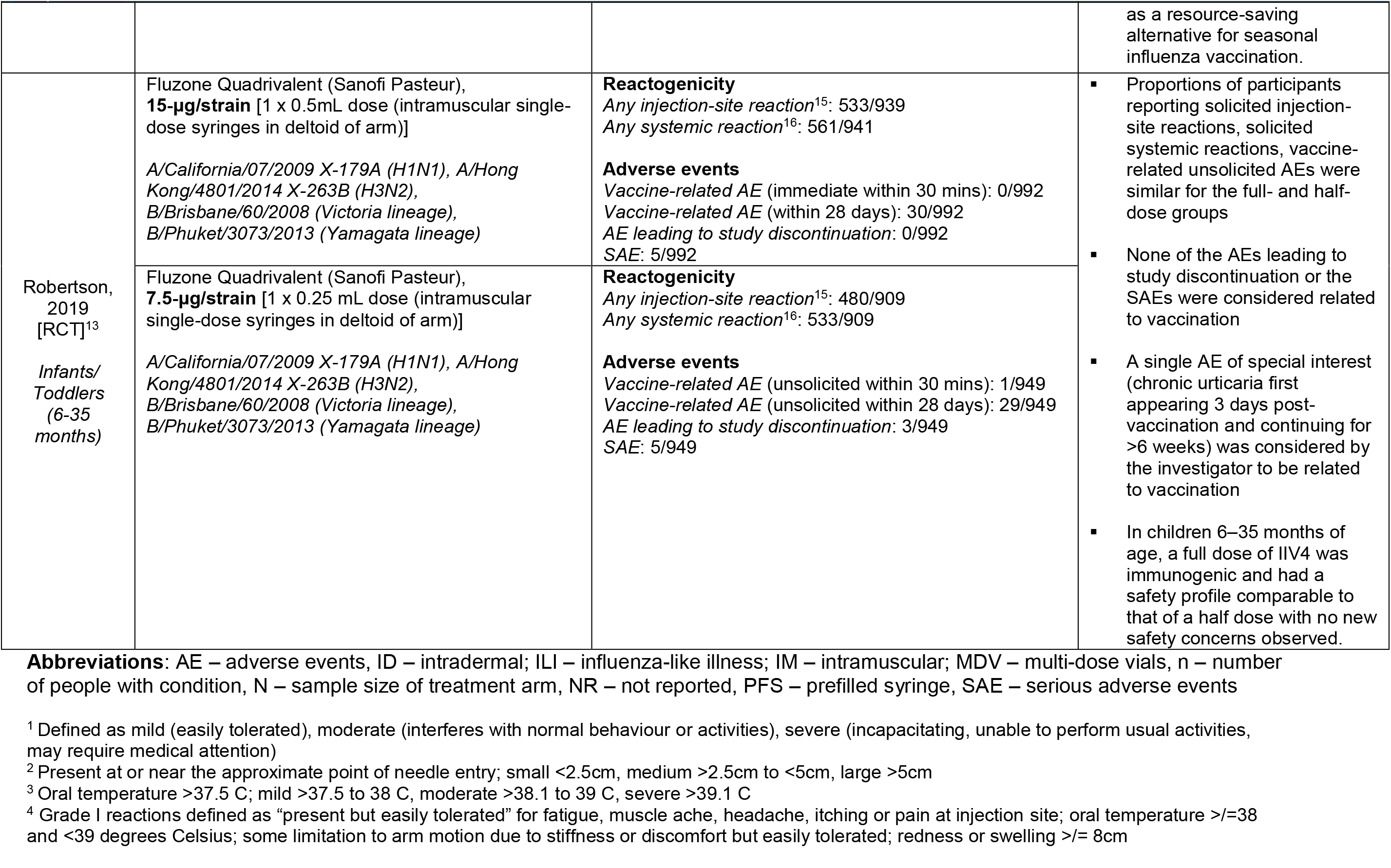
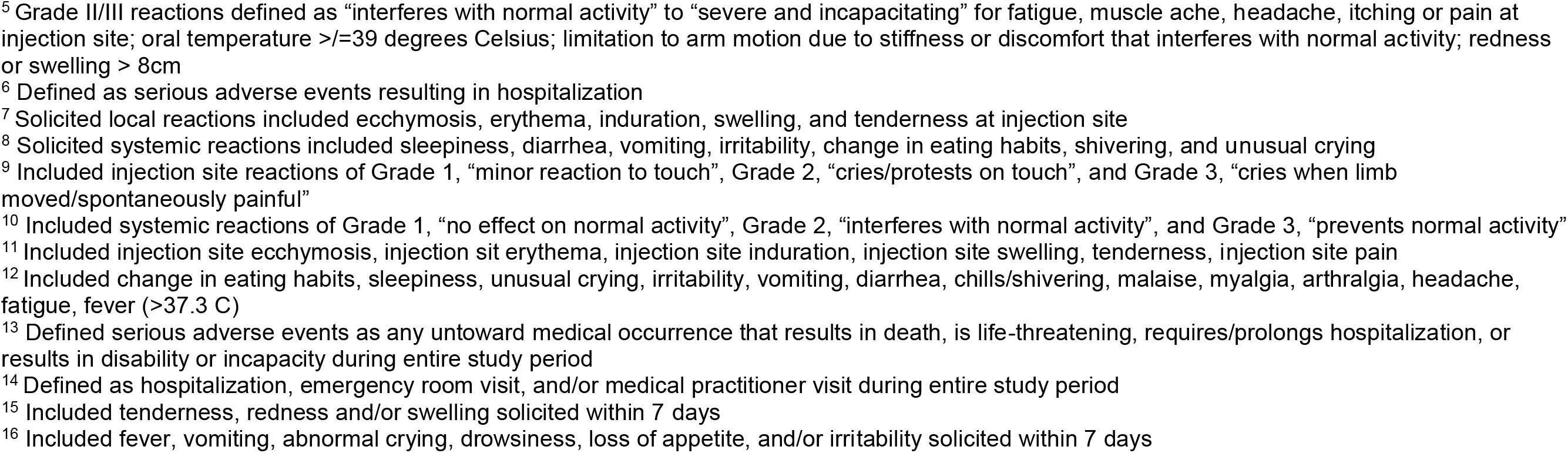

APPENDIX E – Treatment and outcome data
| Author,<br>Year;<br>[Study<br>design]<br>Population | Treatment arms<br>Brand name (manufacturer),<br>HA/strain [dosing (administration)]<br>Included strains | Effectiveness and Safety<br>Outcome (definition):<br>n/N (unless otherwise indicated) | Conclusions |
| --- | --- | --- | --- |
| Kramer,<br>2006<br>[RCT]<sup>14</sup><br>Adults and<br>Seniors<br>(>18 years) | Fluzone (Aventis Pasteur),<br>**15-µg/strain** [1 x 0.5mL dose (Intramuscular into the deltoid region)]<br>*A/Wyoming/3/2003 (H3N2), A/New Caledonia/20/99 (H1N1), and a new B strain, B/Jiangsu/10/2003* | **Effectiveness**<br>*Lab confirmed influenza* (Laboratory confirmation of influenza diagnosis was sought in participants reporting a clinical diagnosis by their physicians): 1/222<br>*Influenza like illness* (Clinical diagnosis of influenza. Participants self-reported four or more symptoms consistent with influenza-like illness (i.e., headache, extreme tiredness, dry cough, fever, muscle or body aches)): 8/222 | There was no significant difference between the full-dose and half-dose groups in the diagnosis of influenza or in the proportion of participants self-reporting four or more symptoms consistent with influenza-like illness.<br>No adverse events were noted by participants from either group or reported to the IRB during the course of the study |
|  | Fluzone (Aventis Pasteur),<br>**7.5-µg/strain** [1 x 0.25 mL dose (Intramuscular into the deltoid region)]<br>*A/Wyoming/3/2003 (H3N2), A/New Caledonia/20/99 (H1N1), and a new B strain, B/Jiangsu/10/2004* | **Effectiveness**<br>*Lab confirmed influenza* (Laboratory confirmation of influenza diagnosis was sought in participants reporting a clinical diagnosis by their physicians): 0/222<br>*Influenza like illness* (Clinical diagnosis of influenza. Participants self-reported four or more symptoms consistent with influenza-like illness (i.e., headache, extreme tiredness, dry cough, fever, muscle or body aches)): 15/222 |  |
| Belshe, 2007<br>[RCT]<sup>16</sup><br>Adults<br>(18-49 years) | Fluzone (Sanofi-Pasteur),<br>**15-µg/strain** [1 x 0.5mL dose (Intramuscular in the non-dominant arm)] | **Reactogenicity – injection site**<br>*Pain*<sup>1</sup>: 15/31<br>*Redness*<sup>2</sup>: 8/31<br>*Swelling*<sup>2</sup>: 7/31<br>**Reactogenicity – systemic**<br>*Fever*<sup>3</sup>: 1/31<br>*Headache*<sup>1</sup>: 15/31<br>*Malaise*<sup>1</sup>: 8/31<br>*Myalgia*<sup>1</sup>: 10/31 | Intradermal vaccine induced significantly more local inflammatory response than Intramuscular vaccine (primary comparison of this study was ID vs IM doses) |
|  | Fluzone (Sanofi-Pasteur),<br>**9-µg/strain** [1 x 0.3mL dose (Intramuscular in the non-dominant arm)] | **Reactogenicity – injection site**<br>*Pain*<sup>1</sup>: 11/31<br>*Redness*<sup>2</sup>: 11/31<br>*Swelling*<sup>2</sup>: 4/31<br>**Reactogenicity – systemic**<br>*Fever*<sup>3</sup>: 1/31<br>*Headache*<sup>1</sup>: 6/31 |  |
|  |  | *Malaise*<sup>1</sup>: 8/31<br>*Myalgia*<sup>1</sup>: 6/31 |  |
|  | Fluzone (Sanofi-Pasteur),<br>**6-µg/strain** [1 x 0.2mL dose (Intramuscular in the non-dominant arm)] | **Reactogenicity – injection site**<br>*Pain*<sup>1</sup>: 14/31<br>*Redness*<sup>2</sup>: 9/31<br>*Swelling*<sup>2</sup>: 4/31<br>**Reactogenicity – systemic**<br>*Fever*<sup>3</sup>: 0/31<br>*Headache*<sup>1</sup>: 9/31<br>*Malaise*<sup>1</sup>: 7/31<br>*Myalgia*<sup>1</sup>: 9/31 |  |
|  | Fluzone (Sanofi-Pasteur),<br>**3-µg/strain** [1 x 0.1mL dose (Intramuscular in the non-dominant arm)] | **Reactogenicity – injection site**<br>*Pain*<sup>1</sup>: 15/31<br>*Redness*<sup>2</sup>: 9/31<br>*Swelling*<sup>2</sup>: 7/31<br>**Reactogenicity – systemic**<br>*Fever*<sup>3</sup>: 3/31<br>*Headache*<sup>1</sup>: 8/31<br>*Malaise*<sup>1</sup>: 3/31<br>*Myalgia*<sup>1</sup>: 7/31 |  |
| Engler, 2008<br>[RCT]<sup>15</sup><br>Adults<br>(18-64 years) | Fluzone (Aventis Pasteur),<br>**15-µg/strain** [1 x 0.5mL dose (Intramuscular injection)]<br>*A/H1N1, A/New Caledonia/20/99; A/H3N2, A/Fujian/411/2002; B, B/Shanghai/361/2002* | **Effectiveness**<br>*Influenza like illness* (Influenza-like illness and complications resulting in either inpatient or outpatient medical encounters were compared between dose groups (by age)): 61/632<br>*Hospitalization or Emergency visits*: 0.3%<br>**Reactogenicity – local/injection site**<br>*Any local reactions* (NR): 8.9%<br>*Arm weakness* (NR): 8.3%<br>*Numbness or burning* (NR): 9.7%<br>*Pain* (NR): 5.9%<br>*Redness or swelling* (NR): 13.4%<br>**Reactogenicity – systemic**<br>*Joint and/or muscle pain* (NR): 4.5% | The relative risk of medical visits and hospitalizations for influenza-like illnesses were similar in the half- and full-dose group regardless of age, and there was no evidence of ILI symptom differences by sex or dose during the 21 days after immunizations.<br>Although injection site pain was greater for full vs half dose (19.9% vs 14.4%; p=.01), when analyzed for clinically significant pain levels significant dose-dependent |
|  |  | **Adverse events**<br>SAE: 2/554 | pain differences were not identified. |
|  | Fluzone (Aventis Pasteur),<br>**7.5-µg/strain** [1 x 0.25 mL dose (Intramuscular injection)]<br>A/H1N1, A/New Caledonia/20/99; A/H3N2,<br>A/Fujian/411/2002; B, B/Shanghai/361/2003 | **Effectiveness**<br>*Influenza like illness* (Influenza-like illness and complications resulting in either inpatient or outpatient medical encounters were compared between dose groups (by age): 64/644<br>*Hospitalization or Emergency visits*: 0.2%<br>**Reactogenicity – local/injection site**<br>*Any local reactions* (NR): 7.5%<br>*Arm weakness* (NR): 6.5%<br>*Numbness or burning* (NR): 7.8%<br>*Pain* (NR): 4.6%<br>*Redness or swelling* (NR): 8.6%<br>**Reactogenicity – systemic**<br>*Joint and/or muscle pain* (NR): 2.2%<br>**Adverse events**<br>SAE: 1/556 | Joint and/or muscle pain were significantly different (p=.02 and p=.03, respectively) by dose.<br>No other adverse event differed significantly by dose |
| Chi,<br>2010<br>[RCT]<sup>17</sup><br>Seniors<br>(>65 years) | Fluzone (Sanofi Pasteur),<br>**15-µg/strain** [1 x 0.5mL dose (intramuscular in deltoid of arm)]<br>A/Solomon Islands/3/ 2006 (A/H1N1),<br>A/Wisconsin/67/2005 (A/H3N2), and<br>B/Malaysia/2506/2004 | **Reactogenicity – injection site, N=64**<br>*Arm motion limitation*: 1 (grade I)<sup>4</sup><br>*Itching*: 4 (grade I)<sup>4</sup><br>*Pain*: 7 (grade I)<sup>4</sup><br>*Redness or discoloration*: 9 (grade I)<sup>4</sup><br>*Swelling*: 13 (grade I)<sup>4</sup><br>**Reactogenicity - systemic, N=64**<br>*Chills*: 1 (grade I)<sup>4</sup>, 1 (grade II/III)<sup>5</sup><br>*Fatigue*: 4 (grade I)<sup>4</sup>, 2 (grade II/III)<sup>5</sup><br>*Fever*: 0<br>*General body ache/pain*: 6 (grade I)<sup>4</sup>, 1 (grade II/III)<sup>5</sup><br>*Headache*: 10 (grade I)<sup>4</sup><br>*Nausea*: 3 (grade I)<sup>4</sup>, 1 (grade II/III)<sup>5</sup><br>**Adverse events** | The two SAEs were acute coronary syndrome and appendicitis and neither were judged to be related to influenza vaccination |
|  |  | SAE<sup>6</sup>: 0/64 |  |
|  | Fluzone (Sanofi Pasteur),<br>**9-µg/strain** [1 x 0.3mL dose (intramuscular in deltoid of arm)]<br>*A/Solomon Islands/3/2006 (A/H1N1),*<br>*A/Wisconsin/67/2005 (A/H3N2), and*<br>*B/Malaysia/2506/2004* | **Reactogenicity – injection site, N=64**<br>*Arm motion limitation*: 1 (grade I)<sup>4</sup><br>*Itching*: 5 (grade I)<sup>4</sup><br>*Pain*: 11 (grade I)<sup>4</sup><br>*Redness or discoloration*: 7 (grade I)<sup>4</sup><br>*Swelling*: 4 (grade I)<sup>4</sup><br>**Reactogenicity - systemic, N=64**<br>*Chills*: 1 (grade I)<sup>4</sup>, 1 (grade II/III)<sup>5</sup><br>*Fatigue*: 6 (grade I)<sup>4</sup>, 1 (grade II/III)<sup>5</sup><br>*Fever*: 1 (grade I)<sup>4</sup><br>*General body ache/pain*: 5 (grade I)<sup>4</sup>, 2 (grade II/III)<sup>5</sup><br>*Headache*: 5 (grade I)<sup>4</sup>, 1 (grade II/III)<sup>5</sup><br>*Nausea*: 2 (grade I)<sup>4</sup>, 1 (grade II/III)<sup>5</sup><br>**Adverse events**<br>SAE<sup>6</sup>: 2/64 |  |
| Cioppa,<br>2011<br>[RCT]<sup>5</sup><br>*Infants/*<br>*Toddlers*<br>*(6-36*<br>*months)* | NR - TIV,<br>**7.5-µg/strain** [2 x 0.25mL dose (intramuscular in deltoid of arm (children 24-35 mo of age) or the anterolateral aspect of the thigh (children <24 mo of age) using prefilled syringes)]<br>*A/Brisbane/59/2007 (A/H1N1)-like virus,*<br>*A/Brisbane/10/2007 (A/H3N2)-like virus, and*<br>*B/Florida/4/2006-like virus (of the influenza*<br>*B/Yamagata lineage)*<br>Agrippal - TIV,<br>**15-µg/strain** [2 x 0.5mL dose (intramuscular in deltoid of arm (children 24-35 mo of age) or the anterolateral aspect of the thigh (children <24 mo of age) using prefilled syringes)]<br>*A/Brisbane/59/2007 (A/H1N1)-like virus,*<br>*A/Brisbane/10/2007 (A/H3N2)-like virus, and*<br>*B/Florida/4/2006-like virus (of the influenza*<br>*B/Yamagata lineage)* | **Reactogenicity**<br>*Any local reaction*<sup>7</sup>: 47%<br>*Any systemic reaction*<sup>8</sup>: 68%<br>**Adverse events**<br>AE (solicited/spontaneously reported): 84%<br>SAE: 0/25<br>**Reactogenicity**<br>*Any local reaction*<sup>7</sup>: 59%<br>*Any systemic reaction*<sup>8</sup>: 50%<br>**Adverse events**<br>AE (solicited/spontaneously reported): 82%<br>SAE: 0/22 | Reactogenicity of the 7.5-µg TIV/QIV formulations was slightly lower than for the corresponding 15-µg formulations.<br>The majority of unsolicited AEs were mild or moderate in severity and none of the SAEs was considered to be related to the study vaccine. |
|  | NR - QIV,<br>**7.5-µg/strain** [2 x 0.25mL dose (intramuscular in deltoid of arm (children 24-35 mo of age) or the anterolateral aspect of the thigh (children <24 mo of age) using prefilled syringes)]<br>*A/Brisbane/59/2007 (A/H1N1)-like virus,*<br>*A/Brisbane/10/2007 (A/H3N2)-like virus,*<br>*B/Florida/4/2006-like virus (of the influenza*<br>*B/Yamagata lineage), and B/Malaysia/2506/2004-*<br>*like antigen virus (Victoria lineage)* | **Reactogenicity**<br>*Any local reaction*<sup>7</sup>: 25%<br>*Any systemic reaction*<sup>8</sup>: 50%<br>**Adverse events**<br>AE (solicited/spontaneously reported): 92%<br>SAE: 1/25 |  |
|  | NR - QIV,<br>**15-µg/strain** [2 x 0.5mL dose (intramuscular in deltoid of arm (children 24-35 mo of age) or the anterolateral aspect of the thigh (children <24 mo of age) using prefilled syringes)]<br>*A/Brisbane/59/2007 (A/H1N1)-like virus,*<br>*A/Brisbane/10/2007 (A/H3N2)-like virus,*<br>*B/Florida/4/2006-like virus (of the influenza*<br>*B/Yamagata lineage), and B/Malaysia/2506/2004-*<br>*like antigen virus (Victoria lineage)* | **Reactogenicity**<br>*Any local reaction*<sup>7</sup>: 39%<br>*Any systemic reaction*<sup>8</sup>: 54%<br>**Adverse events**<br>AE (solicited/spontaneously reported): 71%<br>SAE: 1/28 |  |
|  | Vaxigrip pediatric - TIV (Sanofi Pasteur), **7.5-µg/strain** [2 x 0.25mL dose (intramuscular in deltoid of arm (children 24-35 mo of age) or the anterolateral aspect of the thigh (children <24 mo of age) using prefilled syringes)] | **Reactogenicity**<br>*Any local reaction*<sup>7</sup>: 50%<br>*Any systemic reaction*<sup>8</sup>: 46%<br>**Adverse events**<br>AE (solicited/spontaneously reported): 73%<br>SAE: 1/26 |  |
| Skowronski,<br>2011<br>[RCT]<sup>6</sup><br>*Infants/*<br>*Toddlers*<br>*(6-23*<br>*months)* | Vaxigrip (Sanofi-Pasteur),<br>15-µg/strain [**2 x 0.5mL dose** (Intramuscular injection)]<br>*A/Brisbane/10/07 (H3N2); A/Brisbane/59/07 (H1N1);*<br>*and B/Florida/4/06 (Yamagata lineage)* | **Reactogenicity – injection site**<br>*Induration* (NR): 13.7%<br>*Redness* (NR): 22.6%<br>*Swelling* (NR): 15.3%<br>*Tenderness* (NR): 22.6%<br>**Reactogenicity – systemic**<br>*Fever* (>37.5°C): 8.06%<br>*Irritability* (NR): 59.7%<br>*Decreased appetite* (NR): 38.7% | Local reactions generally were less common in infants than toddlers and more common with full doses versus half doses, but none of these differences were significant.<br>One serious adverse event was reported: a toddler in the half dose group was |
|  |  | *Drowsiness* (NR): 39.5%<br>*Sleep disturbance* (NR): 54.8%<br>**Adverse events**<br>SAE: NR | hospitalized with pneumonia 28 days after the first vaccination. The event was deemed unlikely related to the vaccine. |
|  | Vaxigrip (Sanofi-Pasteur), 15-µg/strain [2 x 0.25mL dose (Intramuscular injection)]<br>*A/Brisbane/10/07 (H3N2); A/Brisbane/59/07 (H1N1); and B/Florida/4/06 (Yamagata lineage)* | **Reactogenicity – injection site**<br>*Induration* (NR): 6.3%<br>*Redness* (NR): 20.3%<br>*Swelling* (NR): 8.6%<br>*Tenderness* (NR): 25.8%<br>**Reactogenicity – systemic**<br>*Fever* (>37.5°C): 11.7%<br>*Irritability* (NR): 60.2%<br>*Decreased appetite* (NR): 43%<br>*Drowsiness* (NR): 41.4%<br>*Sleep disturbance* (NR): 50%<br>**Adverse events**<br>SAE: 1/128 | All of the rate differences were significantly below the allowed 10% increase in reactogenicity for the full dose ($p < 0.001$ for infant and combined analyses, $p < .005$ for toddlers).<br>This randomized controlled trial in infants and toddlers shows that compared with 0.25-mL half-dosing, administration of 2 full 0.5-mL doses of trivalent inactivated influenza vaccine can increase antibody response without increasing reactogenicity in previously unimmunized infants aged 6 to 11 months. |
| Langley, 2012 [RCT]<sup>7</sup><br>Infants/Toddlers (6-35 months) | Fluviral F1 (Sanofi-Pasteur), 7.5-µg/strain [1 x 0.25 mL dose (Intramuscularly in the anterolateral part of the thigh (if the participant was less than 12 months) or in the deltoid region of the arm)]<br>*A/Brisbane/59/2007 (H1N1), A/Uruguay/716/2007 (an A/Brisbane/10/2007 [H3N2]-like virus), and B/Florida/4/2006* | **Reactogenicity – injection site**<br>*Pain* (NR): 45/164<br>*Redness* (NR): 49/164<br>*Swelling* (NR): 22/164<br>**Reactogenicity – systemic**<br>*Drowsiness* (NR) – 44/164<br>*Fever* (NR) – 10/164<br>*Irritability* (NR) – 62/164<br>*Loss of appetite* (NR) – 37/164<br>**Adverse events**<br>SAE: 1/164 | Fluviral F1 group had 1 case of pneumonia resolved<br>Fluviral F2 group had 1 case of bronchial hyper-reactivity in resolving stage<br>The 0.5-mL dose of the study vaccine, when administered to children aged 6–35 months, resulted in a modest but not statistically significant improvement in |
|  |  | *Unsolicited adverse events* (NR): 108/164<br>*Medically attended events* (NR): 52/164 | immunogenicity with clinically similar safety and reactogenicity compared with the 0.25-mL dose. |
|  | Fluviral F2 (Sanofi-Pasteur),<br>**15-µg/strain** [1 x 0.5mL dose (Intramuscularly in the anterolateral part of the thigh (if the subject was less than 12 months) or in the deltoid region of the arm)]<br>*A/Brisbane/59/2007 (H1N1), A/Uruguay/716/2007 (an A/Brisbane/10/2007 [H3N2]-like virus), and B/Florida/4/2006* | **Reactogenicity – injection site**<br>*Pain* (NR): 55/167<br>*Redness* (NR): 54/167<br>*Swelling* (NR): 24/167<br>**Reactogenicity – systemic**<br>*Drowsiness* (NR) – 52/167<br>*Fever* (NR) – 6/167<br>*Irritability* (NR) – 69/167<br>*Loss of appetite* (NR) – 43/167<br>**Adverse events**<br>*SAE*: 1/167<br>*Unsolicited adverse events* (NR): 112/167<br>*Medically attended events* (NR): 40/167 |  |
|  | Vaxigrip (Sanofi-Pasteur),<br>**7.5-µg/strain** [1 x 0.25 mL dose (Intramuscularly in the anterolateral part of the thigh (if the participant was less than 12 months) or in the deltoid region of the arm)]<br>*A/Brisbane/59/2007 (H1N1), A/Uruguay/716/2007 (an A/Brisbane/10/2007 [H3N2]-like virus), and B/Florida/4/2006* | **Reactogenicity – injection site**<br>*Pain* (NR): 17/43<br>*Redness* (NR): 13/43<br>*Swelling* (NR): 5/43<br>**Reactogenicity – systemic**<br>*Drowsiness* (NR) – 11/43<br>*Fever* (NR) – 2/43<br>*Irritability* (NR) – 15/43<br>*Loss of appetite* (NR) – 9/43<br>**Adverse events**<br>*SAE*: NR/43<br>*Unsolicited adverse events* (NR): 24/43<br>*Medically attended events* (NR): 9/43 |  |
| Pavia-Ruz,<br>2013<br>[RCT]<sup>8</sup><br>*Infants/*<br>*Toddlers* | Fluarix (GSK),<br>**15-µg/strain** [1 x 0.5mL dose (intramuscular injection into the right deltoid muscle or anterolateral thigh)] | **Reactogenicity – injection site**<br>*Any injection site reactions*<sup>9</sup>: 514/1086<br>*Pain*: 406/1086<br>*Redness*: 249/1086<br>*Swelling*: 170/1086 | The reactogenicity and safety profile of the study vaccine did not appear to be affected by doubling the dose. |
| (6-35<br>months) | A/Brisbane/59/2007 (H1N1), A/Uruguay/716/2007 (H3N2) and B/Brisbane/3/2007 | **Reactogenicity – systemic**<br>*Any general reactions*<sup>10</sup>: 575/1086<br>*Drowsiness*: 317/1086<br>*Fever*: 69/1086<br>*Irritability*: 387/1086<br>*Loss of appetite*: 273/1086<br>**Adverse events**<br>*Any AE*: 729/1086<br>*SAE*: 29/1086 | One subject in the Flu-15µg group had two SAEs, (apnea and cyanosis) which were considered by the investigator to be possibly related to vaccination. The participant was hospitalized and the events resolved on the same day as they occurred. |
|  | Fluarix (GSK),<br>**7.5-µg/strain** [1 x 0.25 mL dose (intramuscular injection into the right deltoid muscle or anterolateral thigh)]<br>A/Brisbane/59/2007 (H1N1), A/Uruguay/716/2007 (H3N2) and B/Brisbane/3/2007 | **Reactogenicity – injection site**<br>*Any injection site reactions*<sup>9</sup>: 492/1081<br>*Pain*: 403/1081<br>*Redness*: 259/1081<br>*Swelling*: 152/1081<br>**Reactogenicity – systemic**<br>*Any general reactions*<sup>10</sup>: 598/1081<br>*Drowsiness*: 293/1081<br>*Fever*: 67/1081<br>*Irritability*: 386/1081<br>*Loss of appetite*: 281/1081<br>**Adverse events**<br>*Any AE*: 724/1081<br>*SAE*: 35/1081 |  |
|  | Fluzone (Sanofi-Pasteur),<br>**7.5-µg/strain** [1 x 0.25 mL dose (intramuscular injection into the right deltoid muscle or anterolateral thigh)]<br>A/Brisbane/59/2007 (H1N1), A/Uruguay/716/2007 (H3N2) and B/Florida/4/2006 | **Reactogenicity – injection site**<br>*Any injection site reactions*<sup>9</sup>: 467/1090<br>*Pain*: 363/1090<br>*Redness*: 253/1090<br>*Swelling*: 129/1090<br>**Reactogenicity – systemic**<br>*Any general reactions*<sup>10</sup>: 592/1090<br>*Drowsiness*: 298/1090<br>*Irritability*: 375/1090<br>*Fever*: 72/1090<br>*Loss of appetite*: 270/1090 |  |
|  |  | **Adverse events**<br>Any AE: 722/1090<br>SAE: 31/1090 |  |
| Halasa,<br>2015<br>[RCT]<sup>9</sup><br>Infants/<br>Toddlers<br>(6-35<br>months) | Fluzone (Sanofi Pasteur),<br>**7.5-µg/strain** [1 x 0.25 mL dose (intramuscular)]<br>*A/California/7/09 (H1N1)-like virus, A/Perth/16/2009 (H3N2)-like virus, and B/Brisbane/ 60/2008-like virus* | **Reactogenicity**<br>*Redness at injection site:* 8/48<br>*Fever (temperature >39°C after the first dose):* 7/80 | No significant differences between the full-dose or half-dose groups for either the fully primed or naive cohorts for systemic reactions or local reactions when both seasons were combined.<br>The only significant difference in the 2011–2012 season was that 8 of 48 (16.7%) participants in the half-dose group compared with 32 of 96 (33.3%) in the full-dose group had increased redness at the injection site ($P < .05$).<br>No significant differences between the groups in unsolicited AEs, serious adverse events (SAEs), or onset of chronic medical conditions between the dose groups in either the naive or fully primed cohorts, and none of the SAEs were deemed related to the vaccine. |
|  | Fluzone (Sanofi Pasteur),<br>**15-µg/strain** [1 x 0.5 mL dose (intramuscular)]<br>*A/California/7/09 (H1N1)-like virus, A/Perth/16/2009 (H3N2)-like virus, and B/Brisbane/ 60/2008-like virus* | **Reactogenicity**<br>*Redness at injection site:* 32/96<br>*Fever (temperature >39°C after the first dose):* 19/161 |  |
| Phung, 2016<br>[RCT]<sup>10</sup><br>Infants/<br>Toddlers<br>(6-35<br>months) | FLUAD (NR),<br>NR [**1 x 0.5mL dose** (Intramuscular injection)]<br>*A/H1N1, A/H3N2, Strain B* | **Reactogenicity**<br>*Any local reaction*<sup>11</sup>: 45/61<br>*Any systemic reaction*<sup>12</sup>: 36/61<br>**Adverse events**<br>SAE (based on MedDRA v 17.1 definition): 2/61 |  |
|  | FLUAD (NR),<br>NR [**1 x 0.25 mL dose** (Intramuscular injection)] | **Reactogenicity**<br>*Any local reaction*<sup>11</sup>: 63/75 |  |
|  | A/H1N1, A/H3N2, Strain B | Any systemic reaction<sup>12</sup>: 42/75<br>**Adverse events**<br>SAE (based on MedDRA v 17.1 definition): 2/75 |  |
|  | Agrippal S1 (NR),<br>NR [1 x 0.5mL dose (Intramuscular injection)]<br>A/H1N1, A/H3N2, Strain B | **Reactogenicity**<br>Any local reaction<sup>11</sup>: 42/51<br>Any systemic reaction<sup>12</sup>: 24/51<br>**Adverse events**<br>SAE (based on MedDRA v 17.1 definition): 0/51 |  |
|  | Agrippal S1 (NR),<br>NR [1 x 0.25mL dose (Intramuscular injection)]<br>A/H1N1, A/H3N2, Strain B | **Reactogenicity**<br>Any local reaction<sup>11</sup>: 6/10<br>Any systemic reaction<sup>12</sup>: 5/10<br>**Adverse events**<br>SAE (based on MedDRA v 17.1): 0/10 |  |
| Jain,<br>2017<br>[RCT]<sup>11</sup><br>Infants/<br>Toddlers<br>(6-35<br>months) | Flulaval Quadrivalent (GSK),<br>**15-µg/strain** [1 x 0.5mL dose (intramuscular in<br>deltoid region)]<br>A/California/7/2009 (A/H1N1), A/Texas/50/2012<br>(A/H3N2), B/Brisbane/60/2008 (B/Victoria), and<br>B/Massachusetts/2/2012 (B/Yamagata) | **Reactogenicity – injection site (within 7 days)**<br>Pain: 44.0%<br>Redness: 1.4%<br>Swelling: 1.0%<br>**Reactogenicity – systemic (within 7 days)**<br>Drowsiness: 40.6%<br>Fever ($\geq 38.0^{\circ}\text{C}$): 7.9%<br>Irritability/fussiness: 54.4%<br>Loss of appetite: 33.7%<br>**Adverse events**<br>Any AE: 45.5%<br>Vaccine-related AE: 5.9%<br>Any SAE<sup>13</sup>: 1.8%<br>Febrile seizures: 0.4%<br>Medically attended event<sup>14</sup>: 60.2% | None of the febrile seizures or the SAEs were considered by the investigator to be related to vaccination<br>Double-dose IIV4 may improve protection against influenza B in some young children and simplifies annual influenza vaccination by allowing the same vaccine dose to be used for all eligible children and adults. |
|  | Fluzone Quadrivalent (Sanofi Pasteur),<br>**7.5-µg/strain** [1 x 0.25 mL dose (intramuscular in<br>deltoid region)] | **Reactogenicity – injection site (within 7 days)**<br>Pain: 40.1%<br>Redness: 1.4%<br>Swelling: 0.4%<br>**Reactogenicity – systemic (within 7 days)** |  |
| | A/California/7/2009 (A/H1N1), A/Texas/50/2012 (A/H3N2), B/Brisbane/60/2008 (B/Victoria), and B/Massachusetts/2/2012 (B/Yamagata) | Drowsiness: 40.9%<br>Fever ($\geq 38.0^{\circ}\text{C}$): 7.5%<br>Irritability/fussiness: 50.5%<br>Loss of appetite: 33.4%<br>**Adverse events**<br>Any AE: 44.1%<br>Vaccine-related AE: 5.8%<br>Any SAE<sup>13</sup>: 1.7%<br>Febrile seizures: 0.3%<br>Medically attended event<sup>14</sup>: 59.1% | |
| Ojeda.<br>2019<br>[RCT]<sup>12</sup><br>Infants/<br>Toddlers and<br>Children<br>(6 months –<br>17 years) | Vaxigrip Tetra (Sanofi Pasteur) – **PFS**,<br>15-$\mu\text{g}$/strain [1 x 0.5mL dose (intramuscular or deep<br>subcutaneous injection)]<br>A/Michigan/45/2015 (H1N1)pdm09-like virus,<br>A/Hong Kong/4801/2014 (H3N2)-like virus,<br>/Brisbane/60/2008-like virus (B/Victoria lineage), and<br>B/Phuket/3073/2013 (B/Yamagata lineage) | **Reactogenicity, N=142**<br>Any injection-site reaction (solicited within 7 days): 26 (6-35mo), 16 (3-8yr), 42 (9-7yr)<br>Any systemic reaction (solicited within 7 days): 25 (6-35mo), 15 (3-8yr), 35 (9-7yr)<br>**Adverse events, N=147**<br>AE (immediate unsolicited): 1 (9-17 years)<br>Non-serious AE: 25 (6-35mo), 9 (3-8yr), 8 (9-7yr)<br>Vaccine-related non-serious AE: 1 (9-17 years)<br>AE leading to study discontinuation: 0<br>SAE: 0 | Solicited reactions were mostly grade 1 (mild) in intensity and resolved within 3 days.<br>Solicited systemic reactions were reported in more infants aged 6 – 35 months in the MDV group than in the PFS group however, because the 95% CIs were overlapping, this was not thought clinically significant.<br>None of these unsolicited AEs were considered related to a study vaccine by the investigators.<br>There were no differences in reactogenicity or safety between the two vaccine formats. These results showed that the MDV format of QIV was as safe and immunogenic as the PFS format in infants, children, and adolescents. These findings support the use of MDV QIV |
| | Vaxigrip Tetra (Sanofi Pasteur) - **MDV**, 15-$\mu\text{g}$/strain [1 x 0.5mL dose (intramuscular or deep subcutaneous injection)]<br>A/Michigan/45/2015 (H1N1)pdm09-like virus,<br>A/Hong Kong/4801/2014 (H3N2)-like virus,<br>/Brisbane/60/2008-like virus (B/Victoria lineage), and<br>B/Phuket/3073/2013 (B/Yamagata lineage) | **Reactogenicity, N=139**<br>Any injection-site reaction(solicited within 7 days): 27 (6-35mo), 16 (3-8yr), 26 (9-7yr)<br>Any systemic reaction(solicited within 7 days): 33 (6-35mo), 13 (3-8yr), 30 (9-7yr)<br>**Adverse events, N=150**<br>AE (immediate unsolicited): 0<br>Non-serious AE: 31 (6-35mo), 14 (3-8yr), 5 (9-7yr)<br>Vaccine-related non-serious AE: 0<br>AE leading to study discontinuation: 0<br>SAE: 0 | |
|  |  |  | as a resource-saving alternative for seasonal influenza vaccination. |
| Robertson,<br>2019<br>[RCT]<sup>13</sup><br>Infants/<br>Toddlers<br>(6-35<br>months) | Fluzone Quadrivalent (Sanofi Pasteur),<br>**15-µg/strain** [1 x 0.5mL dose (intramuscular single-dose syringes in deltoid of arm)]<br>*A/California/07/2009 X-179A (H1N1), A/Hong Kong/4801/2014 X-263B (H3N2), B/Brisbane/60/2008 (Victoria lineage), B/Phuket/3073/2013 (Yamagata lineage)* | **Reactogenicity**<br>*Any injection-site reaction*<sup>15</sup>: 533/939<br>*Any systemic reaction*<sup>16</sup>: 561/941<br>**Adverse events**<br>*Vaccine-related AE* (immediate within 30 mins): 0/992<br>*Vaccine-related AE* (within 28 days): 30/992<br>*AE leading to study discontinuation*: 0/992<br>SAE: 5/992 | Proportions of participants reporting solicited injection-site reactions, solicited systemic reactions, vaccine-related unsolicited AEs were similar for the full- and half-dose groups<br>None of the AEs leading to study discontinuation or the SAEs were considered related to vaccination<br>A single AE of special interest (chronic urticaria first appearing 3 days post-vaccination and continuing for >6 weeks) was considered by the investigator to be related to vaccination<br>In children 6–35 months of age, a full dose of IIV4 was immunogenic and had a safety profile comparable to that of a half dose with no new safety concerns observed. |
|  | Fluzone Quadrivalent (Sanofi Pasteur),<br>**7.5-µg/strain** [1 x 0.25 mL dose (intramuscular single-dose syringes in deltoid of arm)]<br>*A/California/07/2009 X-179A (H1N1), A/Hong Kong/4801/2014 X-263B (H3N2), B/Brisbane/60/2008 (Victoria lineage), B/Phuket/3073/2013 (Yamagata lineage)* | **Reactogenicity**<br>*Any injection-site reaction*<sup>15</sup>: 480/909<br>*Any systemic reaction*<sup>16</sup>: 533/909<br>**Adverse events**<br>*Vaccine-related AE* (unsolicited within 30 mins): 1/949<br>*Vaccine-related AE* (unsolicited within 28 days): 29/949<br>*AE leading to study discontinuation*: 3/949<br>SAE: 5/949 |  |

**Abbreviations:** AE – adverse events, ID – intradermal; ILI – influenza-like illness; IM – intramuscular; MDV – multi-dose vials, n – number of people with condition, N – sample size of treatment arm, NR – not reported, PFS – prefilled syringe, SAE – serious adverse events
<sup>1</sup> Defined as mild (easily tolerated), moderate (interferes with normal behaviour or activities), severe (incapacitating, unable to perform usual activities, may require medical attention)
<sup>2</sup> Present at or near the approximate point of needle entry; small <2.5cm, medium >2.5cm to <5cm, large >5cm
<sup>3</sup> Oral temperature >37.5 C; mild >37.5 to 38 C, moderate >38.1 to 39 C, severe >39.1 C
<sup>4</sup> Grade I reactions defined as “present but easily tolerated” for fatigue, muscle ache, headache, itching or pain at injection site; oral temperature >=38 and <39 degrees Celsius; some limitation to arm motion due to stiffness or discomfort but easily tolerated; redness or swelling >= 8cm
<sup>5</sup> Grade II/III reactions defined as “interferes with normal activity” to “severe and incapacitating” for fatigue, muscle ache, headache, itching or pain at injection site; oral temperature $\geq 39$ degrees Celsius; limitation to arm motion due to stiffness or discomfort that interferes with normal activity; redness or swelling $> 8$ cm
<sup>6</sup> Defined as serious adverse events resulting in hospitalization
<sup>7</sup> Solicited local reactions included ecchymosis, erythema, induration, swelling, and tenderness at injection site
<sup>8</sup> Solicited systemic reactions included sleepiness, diarrhea, vomiting, irritability, change in eating habits, shivering, and unusual crying
<sup>9</sup> Included injection site reactions of Grade 1, “minor reaction to touch”, Grade 2, “cries/protests on touch”, and Grade 3, “cries when limb moved/spontaneously painful”
<sup>10</sup> Included systemic reactions of Grade 1, “no effect on normal activity”, Grade 2, “interferes with normal activity”, and Grade 3, “prevents normal activity”
<sup>11</sup> Included injection site ecchymosis, injection site erythema, injection site induration, injection site swelling, tenderness, injection site pain
<sup>12</sup> Included change in eating habits, sleepiness, unusual crying, irritability, vomiting, diarrhea, chills/shivering, malaise, myalgia, arthralgia, headache, fatigue, fever ( $>37.3$ C)
<sup>13</sup> Defined serious adverse events as any untoward medical occurrence that results in death, is life-threatening, requires/prolongs hospitalization, or results in disability or incapacity during entire study period

